# Identifying high-risk groups for change in weight and body mass index: population cohort of 11 million measurements in 2.3 million adults

**DOI:** 10.1101/2021.01.19.21249898

**Authors:** Michail Katsoulis, Alvina G Lai, Karla Diaz-Ordaz, Manuel Gomes, Laura Pasea, Amitava Banerjee, Spiros Denaxas, Kostas Tsilidis, Pagona Lagiou, Gesthimani Misirli, Krishnan Bhaskaran, Goya Wannamethee, Richard Dobson, Rachel L Batterham, DK Kipourou, R Thomas Lumbers, Nick Wareham, Claudia Langenberg, Harry Hemingway

**Affiliations:** Institute of Health Informatics, University College London, London, UK; Health Data Research UK, University College London, London, UK; Department of Medical Statistics, London School of Hygiene and Tropical Medicine, UK; Department of Applied Health Research, University College London, London, UK; University College London Hospitals NHS Trust, London, UK; Barts Health NHS Trust, The Royal London Hospital, London, UK; Alan Turing Institute, London, UK; Department of Epidemiology and Biostatistics, School of Public Health, Imperial College London, London, UK; Department of Hygiene and Epidemiology, University of Ioannina School of Medicine, Ioannina, Greece; Department of Hygiene, Epidemiology and Medical Statistics, University of Athens Medical School, Athens, Greece; Department of Epidemiology, Harvard School of Public Health, Boston, USA; Hellenic Health Foundation; Department of Non-Communicable Disease Epidemiology, London School of Hygiene & Tropical Medicine, London, UK; Department of Primary Care and Population Health, University College London, UK; Health Data Research UK London, University College London, London, UK; Department of Biostatistics and Health Informatics, Institute of Psychiatry, Psychology and Neuroscience, King’s College London, London, UK; Centre for Obesity Research, University College London, London, United Kingdom; University College London Hospitals Bariatric Centre for Weight Management and Metabolic Surgery, London; National Institute of Health Research, University College London Hospitals Biomedical Research Centre, London, United Kingdom; Department of Non-communicable Disease Epidemiology, London School of Hygiene and Tropical Medicine, UK; MRC Epidemiology Unit, University of Cambridge School of Clinical Medicine, Cambridge, UK

**Keywords:** Body mass index, weight, change, transition, population-based

## Abstract

**Background:** Adult obesity prevention policies, which are largely untargeted, have met with limited success globally. Population groups with the highest risk of weight gain, if they could be reliably identified using readily available information, might benefit from targeted policy. The relative importance of age, sex, ethnicity, geographical region and social deprivation for weight gain is unknown.

**Methods:** We calculated longitudinal changes in BMI over one, five and ten years and investigated transition between BMI categories using 11,187,383 clinically recorded, repeated measures of BMI from population-based electronic health records of 2,328,477 adults in England (1998-2016). The influence of risk factors was tested using logistic regression.

**Findings:** The youngest adult age group (18-24 years) was more strongly associated with risk of weight gain than older age, male sex, socioeconomic deprivation, ethnicity or geographic region. Among the youngest adults, the top quartile gained 15.9kg in men and 12kg in women at 10 years. The odds of transitioning to a higher BMI category over 10 years were 4-6 times higher in the youngest (18-24 years) compared to oldest (65-74 years) individuals; odds ratio (95% confidence interval) 4.22 (3.85-4.62)) from normal weight to overweight or obesity, 4.60 (4.06-5.22) for overweight to obesity, and 5.87 (5.23-6.59) from obesity to severe obesity in multiple adjusted analyses. Among the youngest adults, socially deprived men were at greater risk of transitioning from normal weight to overweight (72%) and from overweight to obesity (68%) over 10 years. We provide an open access online risk calculator (https://pasea.shinyapps.io/bmi_shiny_app/) and present high resolution obesity risk charts over a 1-, 5- and 10-year follow-up.

**Interpretation:** A radical shift in policy is required to focus on those at highest risk of weight gain-young adults-for individual and population level prevention of obesity and its long-term consequences for health and health care.

**Funding:** BHF, HDR-UK, MRC

## INTRODUCTION

Adult obesity prevention policies, which are largely untargeted, have had limited success globally[1,2] and the high prevalence of obesity is predicted to increase dramatically[3,4]. Population-wide approaches to obesity prevention could be complemented by new targeted approaches to intervention if it were possible to identify population groups at highest risk of weight gain using information readily available in national public health systems.[1] Current obesity prevention policies are informed by population evidence from cross-sectional surveys [5], which by definition cannot evaluate weight change, and highlight the higher cross-sectional prevalence of obesity in middle age, socially deprived areas, and in some ethnic groups.[6] In longitudinal research, younger compared to older adults have a greater risk of weight gain in some[7,8] but not all studies [9] (see Table S1.1 in the Appendix)

An emerging data science opportunity to identify population groups at high risk for weight gain comes from population-based electronic health records (EHR). Understanding how adult age, sex, ethnicity, geographical region and social deprivation might combine to identify groups at highest risk of weight gain, is essential if targeted policy is to be considered. We found no previous studies providing such evidence. Population-based electronic health records offer valid measurements of weight and BMI change [10-11] and have potential advantages (compared to consented cohort studies) of sufficient scale to evaluate the joint contribution of several factors, contemporary and updatable and low-cost measurements.

Here we report, for the first time, the use of population-based EHR as a way of answering longstanding calls for ‘monitoring basic population weight data’[12] in the context of weight change. We present within-individual 1-, 5- and 10-year BMI change patterns with the following objectives: (i) to test the extent to which temporal trends in mean BMI 1999-2016 are replicated between electronic health records and health survey data [13]; (ii) to compare the extent and distribution of BMI change across age and within BMI categories; (iii) to estimate the associations of age, sex, social deprivation, ethnicity and region with transitions across BMI categories and (iv) to provide a risk calculator (online and chart), showing how these risk factors combine to identify high risk groups for transitioning to higher BMI categories.

## METHODS

### Study population

We used clinically recorded measures of height, weight and BMI from usual practice in primary care in England in the Clinical Practice Research Datalink. This study was carried out as part of the CALIBER© resource (https://www.ucl.ac.uk/health-informatics/caliber and https://www.caliberresearch.org/). CALIBER, led from the UCL Institute of Health Informatics, is a research resource providing validated electronic health record phenotyping algorithms and tools for national structured data sources. These EHR data came from patients registered in 400 primary care practices and were accessed through the CALIBER programme[14]. We studied 2,328,477 adults with recorded BMI measurements in England between 1998 and 2016 (see Appendix, Section 2). The study was approved by the MHRA (UK) Independent Scientific Advisory Committee [ISAC number: 18_010R], under Section 251 (NHS Social Care Act 2006).

### Weight, height and BMI EHR phenotypes

NHS policy is for weight and height to be recorded at registration with a general practice and then repeated for health checks and at clinical discretion[11]. We extracted height (in meters, m), weight (in kg) and BMI (in kg/m^2^) measurements, as recorded in usual care by physicians, nurses and health care assistants using a range of devices. We included individuals with BMI and weight measurements recorded during the period in which they had eligible linked data, with at least one year of follow-up time. Weight is one of the most frequently recorded and repeated measures in the general population as part of preventive health practice, while BMI has proven to be representative when it is combined with model-based imputation from Health Survey from England[10]. We excluded BMI records (Figure S5.1A) during pregnancy; where two observations on the same day differed by more than 0.5 kg/m^2^; individuals whose highest BMI was more than double their lowest BMI record and observations where the absolute difference between recorded and calculated BMI on the same date was more than 1 kg/m^2^. We defined four categories: underweight (BMI<18.5kg/m^2^), normal weight (BMI≥18.5 and BMI<25kg/m^2^), overweight (BMI≥25 and BMI<30kg/m^2^), non-severe obesity (BMI≥30kg/m^2^ and BMI<40kg/m^2^) and severe obesity (BMI≥40kg/m^2^).

### Demographic factors

We defined age (at the date of the first BMI record) using age 18-24 years as the youngest adult category, sex (men/women), ethnicity [White, Asian (Chinese, Indian, Pakistani, Bangladeshi, other Asian), Black (Black African, Black Caribbean, Black other) and Mixed/Other], an area based measure of social deprivation, index of multiple deprivation (IMD in quintiles from low to high deprived), and region (London, South West, South Central, South East, West Midlands, East of England, central North East, and North West).

## STATISTICAL ANALYSIS

### Comparison of time trends in mean BMI

From the 2,328,477 individuals in CALIBER (England), we selected at random one BMI observation and we compared the mean BMI 1999-2016 in England with the corresponding US trend (reported from NHANES[15] over the same period, by age group. We also compared the EHR measures of BMI with standardised survey measures of BMI from an annual cross sectional survey, the Health Survey for England, by age and sex[13].

### Defining 1, 5 and 10-year within individual BMI change

To estimate the 1, 5 and 10-year BMI change we selected at random one pair of measurements per individual within window intervals of 6 months to 2 years, 4 years to 6 years and 8 years to 12 years respectively. We assumed that any change in BMI for each individual was linear in the window periods (see Appendix, Section 2). We then calculated the 10^th^, 25^th^, 50^th^, 75^th^ and 90^th^ centile of the distribution of the 1-, 5- and 10-year BMI change, by age and BMI category. We calculated weight change for an average height individual (1.76m for men and 1.62m for women). For more details, see Appendix, Section 5.

### Missing values

The problem of missing values arises because not all individuals had two BMI measurements within the specific windows of interest (see figure S2.1B). We assumed that data for BMI change were missing not at random (MNAR) i.e. missing values on BMI change might depend on BMI change itself. For this reason, we applied multiple imputation with delta adjustment, as it provides a flexible and transparent means to impute missing data under MNAR mechanisms[16]. More specifically, we created multiple copies of datasets (N=10) with missing values of BMI change being imputed values sampled from their predictive distribution (under MAR), through multiple imputation. We then make the missing systematically different from the observed by adding a value (denoted by delta) to our imputed BMI change values. These delta values need to be specified, using external data. Here, these were derived from the average 10-year BMI change by age and sex in the population, calculated from the HSE. For more details, see Appendix, section 3.

### Transitions between BMI categories

We calculated the age-standardised transition between BMI categories at 1, 5 and 10 years, based on the age structure of the English population (Office for National Statistics), and BMI information from the survey (for more details, see Appendix, section 6). We produced risk charts estimated the 10-year BMI transition, by age, index of multiple deprivation and current BMI overall, as well as separately for men and women non-parametrically. We also created an online risk calculator (https://pasea.shinyapps.io/bmi_shiny_app/) for the 10-BMI transition based on socio-demographic factors. The estimation of the online calculator is semi-parametric; that is the (non-parametric) 10-year BMI transition by age, sex, index of multiple deprivation and current BMI overall is calculated in white individuals and then multiplied by the relative risk of transitions of ethnicity (from figure 4), after converting the corresponding odds ratios to relative risk (see Appendix, section 3.5).

We estimated the odds ratios of the sociodemographic factors for the transition between BMI category within 1-, 5- and 10-year periods, using logistic regression models. In all these models, we additionally adjusted for BMI at baseline, family history of CVD, use of diuretics and for prevalent chronic conditions at baseline (cancer, diabetes, CVD, psychological conditions, dementia, chronic kidney disease, systemic lupus erythematosus, rheumatoid arthritis, gout, ulcerative colitis, Parkinson disease, multiple sclerosis and renal disease).

### Sensitivity analyses

In the sensitivity analyses, we applied the same analysis, but i) excluded all missing values (complete case analysis), ii) assumed data were missing at random, i.e used standard multiple imputation without delta adjustment, iii) without controlling for the prevalent chronic conditions, iv) excluded individuals with prevalent chronic diseases, and v) excluded individuals with prevalent chronic diseases and those who developed chronic conditions during follow-up (Appendix, Section 5). Definitions of chronic diseases and drugs are in the Appendix (Section 4).

## RESULTS

### Validation between EHR (CALIBER data) and Health Survey for England

Differences in mean BMI estimates between CALIBER (EHR) and HSE data were negligible, especially for individuals aged<65 years old (see Figure S3.3.1 in the Appendix).

### Time trends in mean BMI in England and US

The mean BMI increased in England between 1999-2016 based on EHRs in each age group, and the rate of increase was similar to that in the US (based on consented survey data from NHANES) – with the US mean BMI higher by about 2kg/m^2^ in each biannual period and each age group (Figure 1).

**Figure 1:**
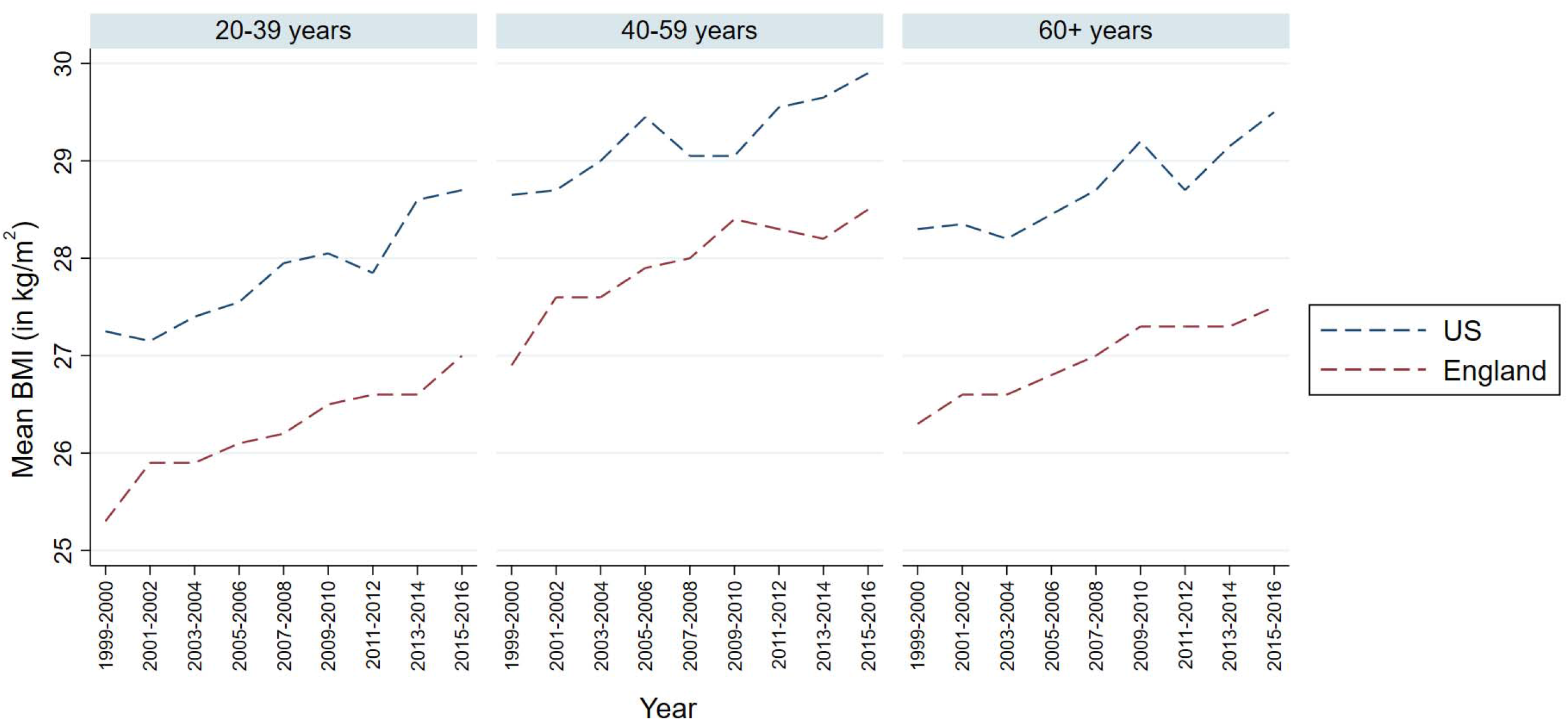
Trends in BMI 1999-2016 by age in England (EHR data from CALIBER, n=2M) and the US (survey data NHANES n=47K)

### Initial and estimated weight at 10 years

We calculated the initial and estimated average of weight after 10 years in CALIBER, by age and sex (figure 2). We observed that younger individuals gained on average more weight compared to older individuals within a decade. For example, the average initial weight of men aged 18-24 was 80.8kg and their average estimated weight after 10 years was 90.2kg.

**Figure 2:**
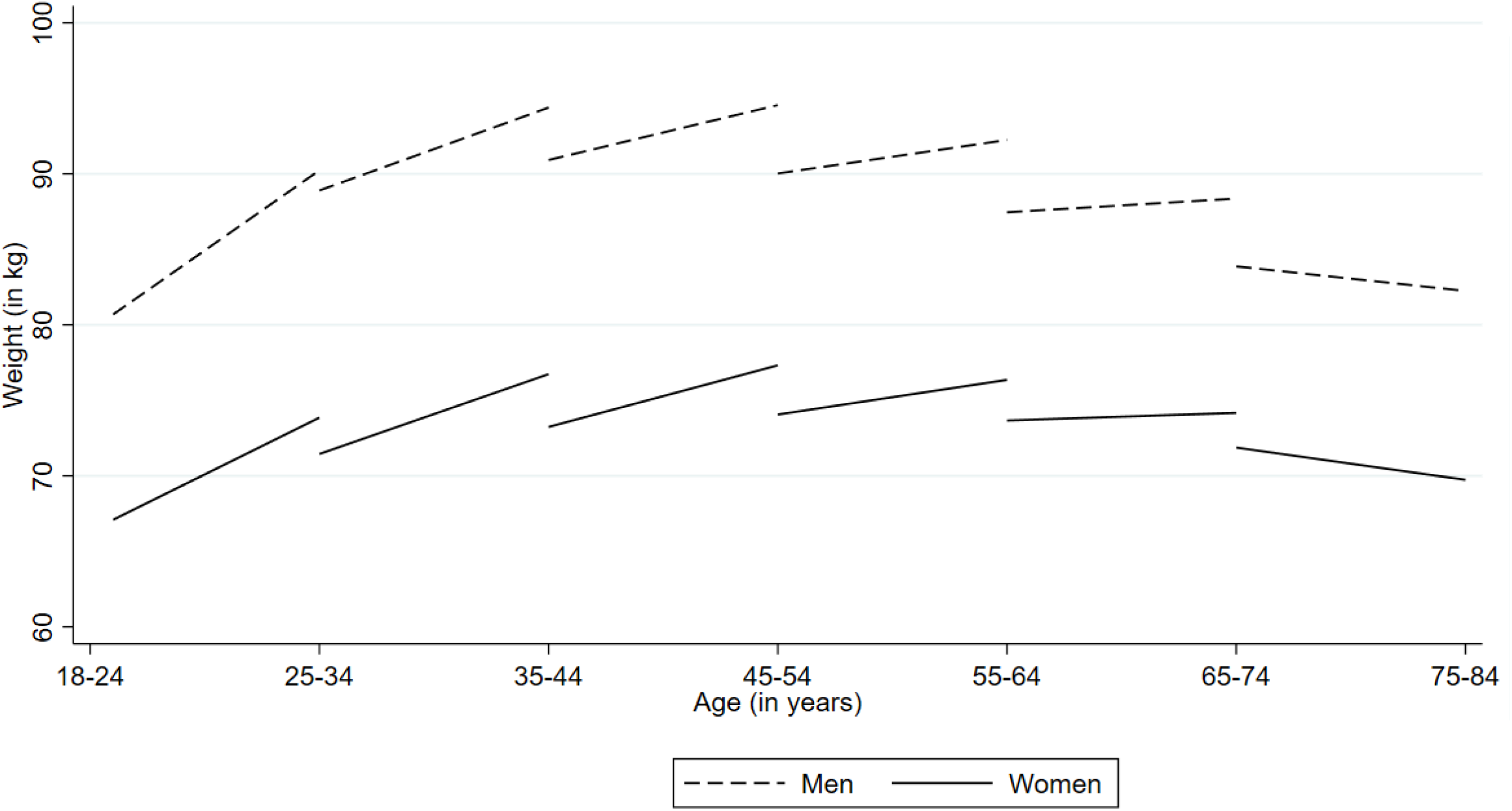
Weight trajectories between 1998 to 2016 in CALIBER. Average^∗^ of initial weight (beginning of each line; measured between 1998 and 2008) and estimated average of weight after 10 years (end of each line), by sex and age (youngest cohort 18-24 years old; oldest cohort: 65-74 years old) ^∗^Estimated averages of weight have been calculated from the complete-case analysis of 10-year weight change

### Age specific distribution of BMI and weight change

The youngest adults (aged 18-24 years) had the highest increase in 10-year BMI (highest median and 90^th^ centile), and the widest distribution (inter-decile range (IDR = 90^th^ centile-10^th^ centile) in each BMI category. Each of these measures decreased progressively with age (Figure S2.2) in each BMI category. For example, among normal weight individuals aged 18-24 years the median and 90^th^ centile of BMI change was 9.9% and 31.3%, compared to 0% and 15% in individuals aged 65-74 years respectively. Younger compared to older adults had wider BMI change distributions among all initial BMI categories. Among initially normal weight individuals those aged 18-24 years (vs 65-74 years) had an inter-decile range (IDR)=41.4% vs 30% respectively.

Figure S2.2 reports the expected weight change for a man and a woman of average height (1.76m and 1.62m respectively). It suggests that the youngest adults had the highest weight change: top quartile of whom at 10 years gained 15.9kg (men) and 12kg (women). The top decile of individuals aged 18-24 years gained 23.4kg (men) and 18.8kg (women) at 10 years. Figures S2.3 and S2.4 show that this finding is consistent across all BMI categories.

### Absolute risk of BMI category transitions

Figure 3 shows that among normal weight individuals, the absolute risk of transitioning to overweight or obesity at one, five and ten years, was 13%, 22.5% and 29%. Among overweight individuals 11%, 20.6% and 28.6% developed obesity at one, five and ten years respectively. Among individuals with obesity, we found that 14.7%, 18.1% and 18.9% transitioned to overweight and 0.6%, 1.6% and 2.8% to normal weight at one, five and ten years respectively.

**Figure 3:**
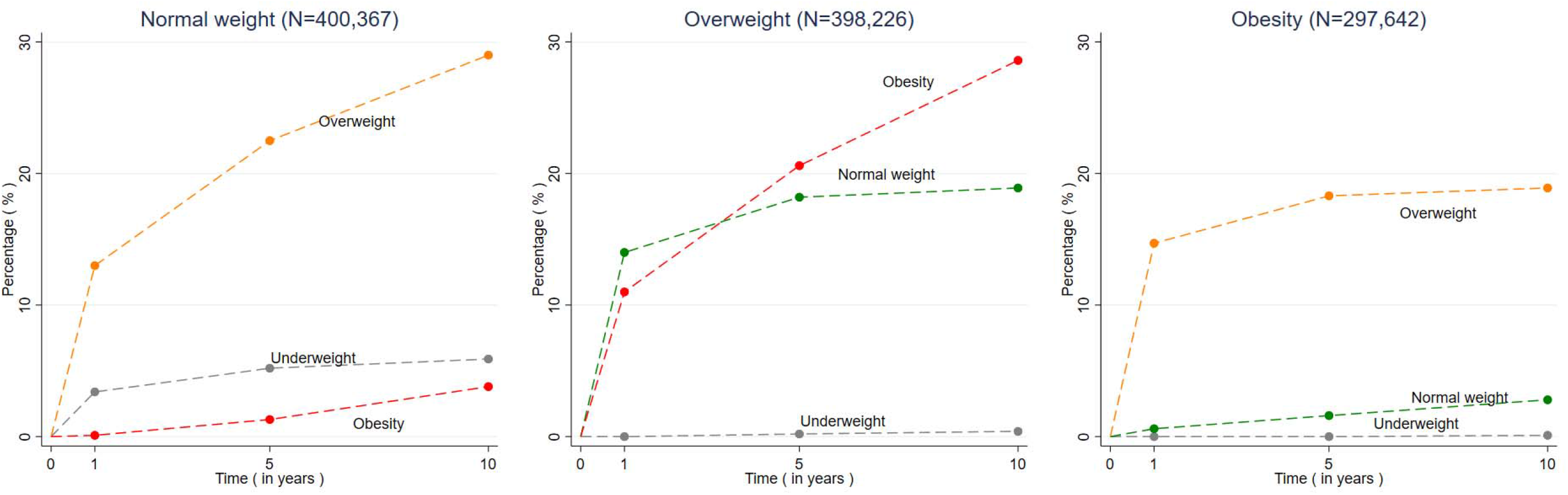
Estimated age-standardised risk of transitioning^†^ to normal weight, overweight and obesity by BMI groups after one, five and ten years of follow-up † The risk of transitioning was standardised population to the English population based on age structure [using information from Office of National Statistics (ONS)] and prevalence of BMI categories (from Health Survey from England (HSE)] between 1998 and 2016. For more details, see Appendix, section 6

### Absolute risk and odds ratios of BMI transitions by socio-demographic factors

Figure 4 shows that the youngest adults, age 18-24 years, had the highest absolute risk (AR=37%) and odds ratio (vs 65-74 years) OR=4.22 (95%CI: 3.85-4.62) of transitioning from normal weight to overweight or obesity at ten years. Other demographic factors were less associated with this transition; more deprived individuals [OR (5^th^ vs 1^st^ quintile of IMD)=1.23 (1.18-1.27) men (vs women) [OR= 1.12 (1.08-1.16)] and black individuals (vs white Caucasians) [OR= 1.13 (1.04-1.24)]. The youngest adults had the highest AR 42% and OR (vs 65-74 years) =4.60 (4.06-5.22)] for transitioning from overweight to obesity. The youngest adults had also the highest AR 22%; and OR (vs 65-74 years) =5.87 (5.23-6.59)] for transitioning from non-severe to severe obesity. Figure S2.5 shows that the association of index of multiple deprivation with transition to higher BMI categories is even more pronounced among the younger adults aged 18-24 years old.

**Figure 4:**
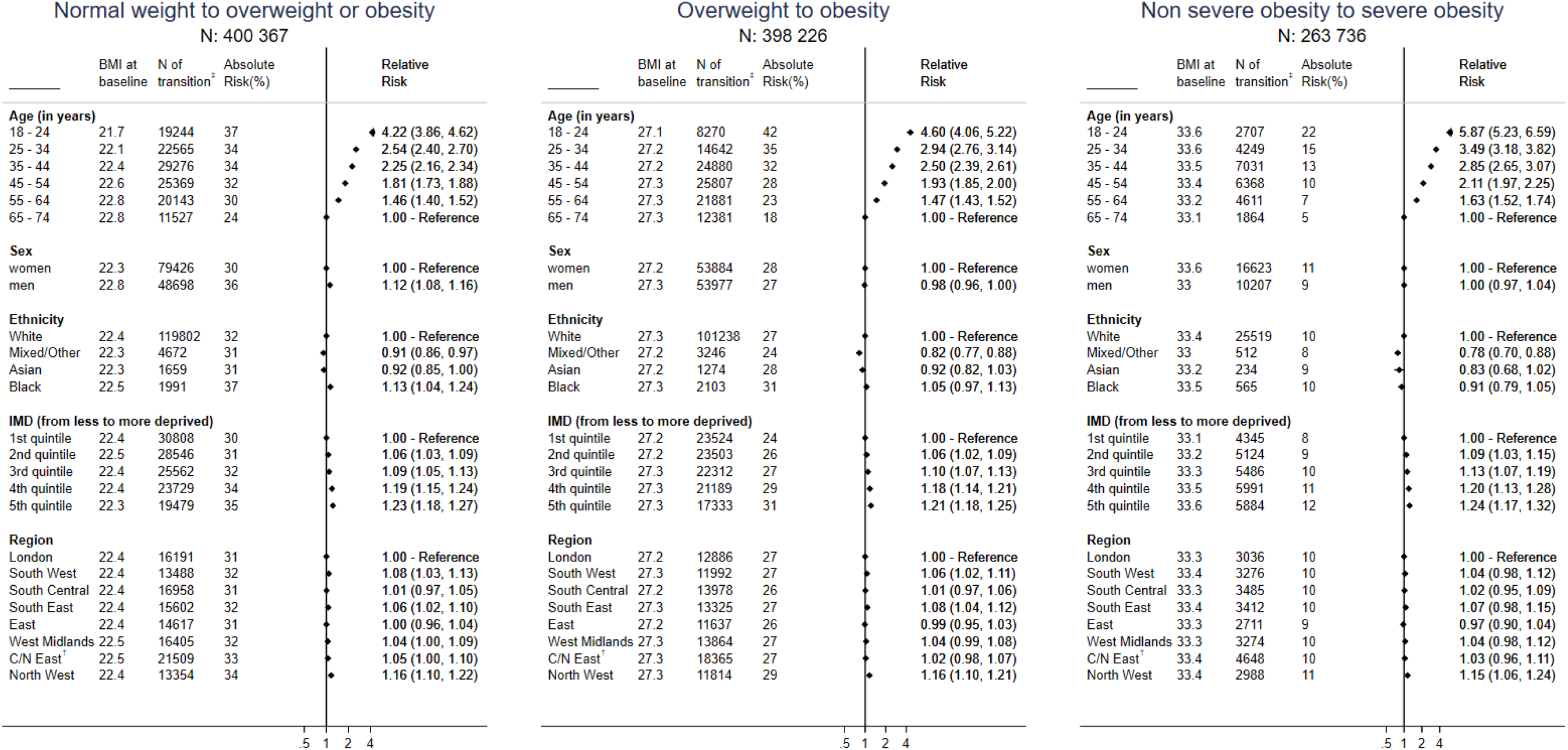
Absolute risks and odds ratios of transitioning at ten years from normal weight to overweight or obesity, from overweight to obesity and from non-severe to severe obesity, by age, sex, ethnicity, social deprivation and region ^∗^Mutually adjusted for BMI (at baseline), age group, gender, Index of multiple deprivation (IMD – quintiles; in categories), ethnicity, region, use of diuretics, prevalence of chronic and mental health diseases †Central/North East ‡N of individuals who transitioned to higher BMI categories

We also considered the risks for remaining ≥30kg/m^2^ (obesity). Figure S2.6 reports that middle-aged individuals 35-54 years had the highest absolute and odds ratio for remaining ≥30kg/m^2^ [for 35-44 years: AR 84% and OR (vs 65-74 years) =2.09 (95%CI 1.97-2.22) and for 45-54 years: AR 83% and OR (vs 65-74 years) =1.97 (95%CI 1.87-2.08)]. Moreover, we observed higher risk for remaining ≥30kg/m^2^ in the more deprived individuals [OR (5^th^ vs 1^st^ quintile of IMD)=1.09 (1.04-1.13)].

We observed similar findings for the youngest adult for the transitions at higher BMI categories at one and five years (see Figures S2.7-8).

### Risk calculator for transitions between weight categories, combining age, sex, social deprivation and initial BMI

We showed that within each strata defined by initial BMI, sex and social deprivation the risks for transitioning to higher BMI categories were substantially higher in the youngest adult age groups. For example, Figure 5 reports that among individuals who are close to the BMI transition cut-offs, the probability of men aged 18-24 years (vs age 65-74 years) living in the median quintile of deprivation (quintile 3) to transition over 10 years from normal weight to overweight was 68% vs 44%, from overweight to obesity was 65% vs 38%, and from non-severe to severe obesity was 47% vs 27%.

**Figure 5:**
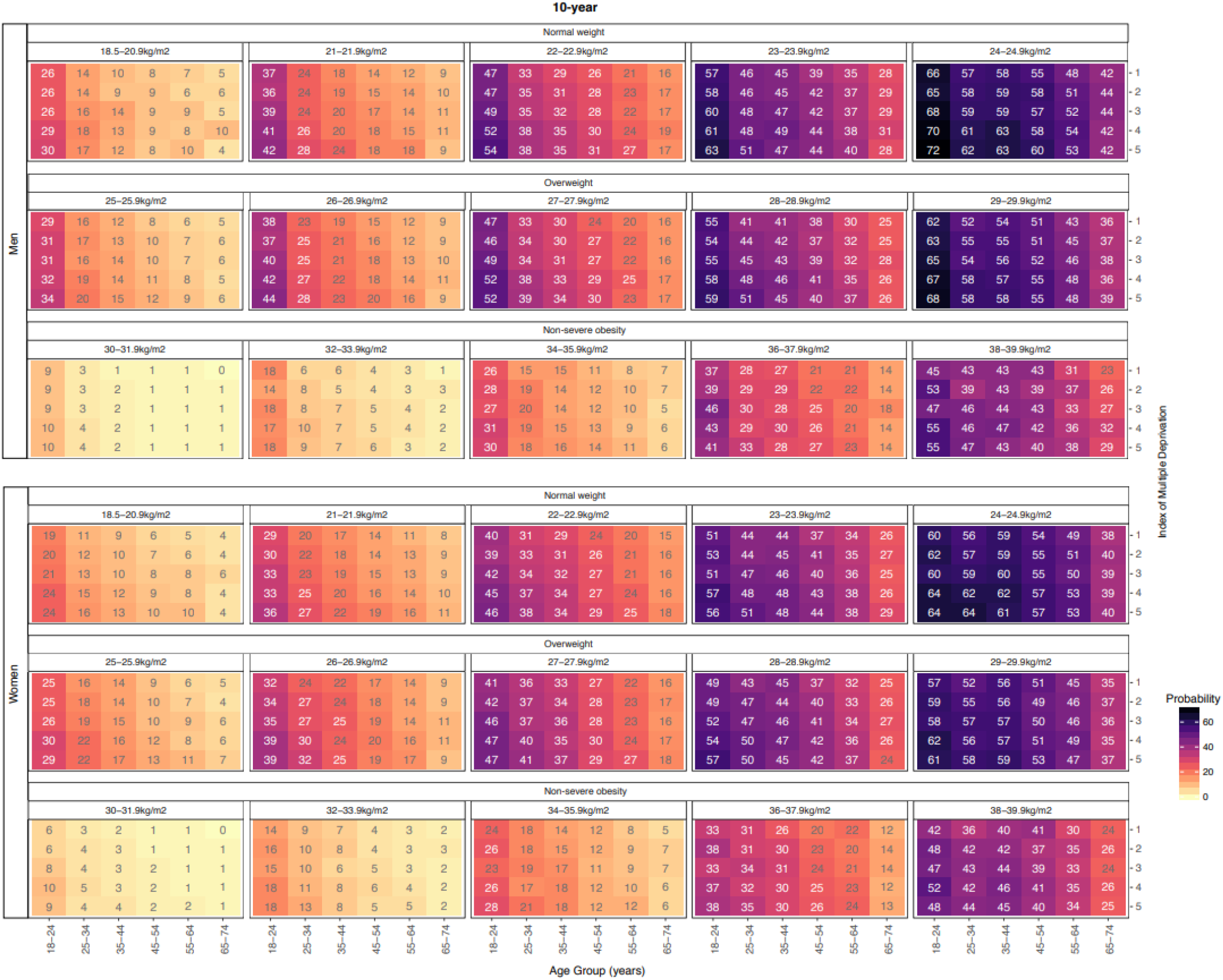
Absolute risk of transitioning to a higher BMI category over 10 years, by initial BMI category, age, sex and social deprivation (using index of multiple deprivation)

By contrast there were only modest modifications of the age association by social deprivation and gender (Figure 5 and https://pasea.shinyapps.io/bmi_shiny_app/). For example, among individuals who are close to the BMI transition cut-offs, the probability of individuals aged 18-24 years living in the most deprived (quintile 5) vs least deprived (quintile 1) areas to transition over 10 years from normal weight to overweight was 72% vs 66%, from overweight to obesity was 68% vs 62% and from non-severe to severe obesity was 55% vs 45% for men. The corresponding figures for women were 64% vs 60%, 61% vs 57% and 48% vs 42% respectively.

We found consistent patterns of age-related transitions to higher BMI categories at 1 year (Figure S2.9) and 5 years (Figure S2.10).

### Sensitivity analysis

Our finding that the youngest adults were at highest absolute and relative risk of BMI transition to a higher BMI category (compared to older adults), with stronger associations than seen for sex, social deprivation, ethnicity, region was robust to each of our sensitivity analyses (see Appendix, Figures S4.1-S4.5).

## DISCUSSION

We found that the youngest adults (18-24 years) were at much greater risk of weight gain than were older adults, men, ethnic minorities or people living in socially deprived neighbourhoods. In by far the largest study to date (see Table S1.1 in the Appendix) we provide the first high resolution obesity risk calculator based on age, sex, social deprivation, ethnicity and current BMI to identify within-person change in BMI, and transition across obesity categories. Our findings demonstrate that it is possible to identify those at highest risk of weight gain using EHR and information readily available to public health agencies, and that young adults should be a major focus of strategies to prevent the onset of overweight and obesity.

### EHR data

We demonstrate the value of new approaches to understanding patterns of weight gain using population based EHR, which offer large scale, but largely untapped sources of information. Previous studies have shown the validity of single measures of BMI recorded in primary care, [10-11] and of BMI change in people with obesity[17] or after bariatric surgery.[18] We extend these studies to a population-wide approach on BMI change, providing new evidence of validity of EHR data by demonstrating the comparability of temporal trends in BMI comparing large scale EHR with small scale survey data. Furthermore, we replicated (and extended) previous findings from smaller consented longitudinal studies (see Appendix, Table S1.1). We propose that public health agencies ‘monitoring basic population weight data[12] should incorporate within person change information from EHR, which are increasingly available in the US and other countries.[19,20]

### Youngest adults gain most weight: middle age remain in obesity

The youngest adults (age 18-24 years) were at highest absolute and relative risk to transition from normal weight to overweight or obesity, from overweight to obesity, and from non-severe to severe obesity at 1-, 5- and 10-years. Furthermore, individuals aged 35-54 years had the highest risk for remaining in obesity (see Appendix, Figure S2.6), further emphasising the importance of early intervention.

### Policy implications

Our findings have policy implications in three inter-related areas: obesity prevention, the health priorities of young adults and addressing lockdown associated weight gain. First, obesity prevention policies should identify groups at high risk of weight gain for targeted intervention. Young adults may offer particular opportunities for intervention because of changes in dietary and exercise behaviours [21], relationships[22], higher education and employment status [23] which may influence weight gain. For example, BMI may increase in women and men in the course of cohabiting relationships and marriage [22].

Identifying groups at the highest risk of weight gain is not currently recommended in obesity prevention policies[13,24] and is not mentioned in reports on ‘new thinking’ in obesity prevention.[1] Our findings suggest that regional and national initiatives aimed at socially deprived areas, might be complemented by targeting young adults. Secondly, young adulthood is increasingly a focus of health policy seeking to address a wide range of issues including substance abuse, sexual health, mental health, violence and injuries, and criminality [25]. An extensive review of preventive interventions in young adulthood made no mention of the fact that young adults are at highest risk of weight gain and found ‘no programs addressing overweight and obesity and healthy living were identified[26]. From both perspectives-obesity prevention, and health in young adulthood –it is important to address the challenge that (with the exception of pregnancy complications), most of the adverse health consequences of obesity (including certain cancers, cardiovascular diseases, type II diabetes, severe COVID-19 infection) occur decades later in middle and older ages. Thirdly, repeated lockdowns during the COVID-19 pandemic, can lead to reduced levels of physical activity and energy expenditure in young adults[27] with adverse effect on weight gain [28].

### Strengths and limitations

Population-based EHR are the only high-resolution source of information on BMI change with sufficient sample size to estimate risk across combinations of age, sex, ethnicity, index of multiple deprivation and BMI category. This was the first large-scale study (2.3M of individuals – 11M of BMI measurements) to focus on modern BMI change patterns in a high resolution at one, five and ten years and its socioeconomic and demographic factors.

Our study had some limitations. Our focus was on the 1-, 5- and 10-year BMI change; nevertheless, individuals measured their bodyweight in their practice infrequently [11]. To address this problem, we considered window periods within which individuals had a pair of BMI measurements (see Appendix Section 2). We assumed linear BMI change in these windows. Despite the large sample size of our study, it is plausible to assume that our sample was not representative for calculating BMI change, as the individuals who measured their bodyweight multiple times in the practice had higher prevalence of chronic disease. To account for this limitation, we applied multiple imputation with delta adjustment (to account for missingness not at random) [16], using information from mean BMI change from HSE. We additionally applied extensive sensitivity analysis and found the main findings to be robust to alternative assumptions about the missing data. However, we cannot exclude the possibility that we mis-specified the imputation models, even if it is unlikely that the resulting bias would be major. Moreover, we utilised observational data, so it is possible that some of the health variables we used, both in the imputation model, as well as in Figure 4, were mis-measured. Finally, we didn’t use different BMI cut-offs for Asian individuals to avoid computational complexity, nevertheless the proportion of Asian individuals is very small in this study (<2%) and did not affect our results, as well as the main conclusion derived from this project (i.e. that younger age is the most important factor for BMI gain).

### Research implications

Our findings suggest the importance of intervention research questions at both the population and individual level. First, at the population level, are interventions developed for, and targeted at, groups identified at highest risk of weight gain effective at achieving weight maintenance (in the normal weight) and preventing weight gain? Given the childhood and adolescent trajectories of weight gain, what are the complementary approaches before and after age 18 years[29]? Second, at the individual level is it possible to communicate risk of weight gain information and incorporate into interventions designed to affect youngest adults’ knowledge, attitude and behaviour. Individuals may have, or seek, additional and actionable predictive information (e.g. accelerometery data) which may inform the design of interventions. Further, to what extent is there a role for primary care professionals[30] and other clinicians in using and recording, risk of weight gain information, alongside other available EHR information.

## Conclusion

Adults aged 18-24 years are at highest absolute and relative risk for transitioning to higher BMI categories; there are smaller additional contributions to risk of living in socially deprived areas and ethnicity. Young adulthood offers an important, currently neglected, opportunity for both population prevention of the onset of obesity and identification of individuals at high risk of weight gain for targeted interventions.

## Supporting information

STROBE

## Data Availability

Data are not available

## ACKNOWLEGDEMENTS

We thank Bianca DeStavola for her comments especially regarding addressing the missing values.

This study is based in part on data from the Clinical Practice Research Datalink obtained under license from the UK Medicines and Healthcare products Regulatory Agency. The data is provided by patients and collected by the NHS as part of their care and support. The interpretation and conclusions contained in this study are those of the author/s alone

## CONTRIBUTION

Research question: MK, CL, HH Funding: MK, SD, HH

Study design and analysis plan: MK, AL, KDO, MG, KB, CL, HH

Preparation of data, including electronic health record phenotyping in the CALIBER open portal: MK, SD

Statistical analysis: MK, MG

Drafting initial versions of manuscript: MK, CL, HH

Drafting final versions of manuscript: MK, CL, HH

Critical review of early and final versions of manuscript: All authors

## FUNDING

MK is funded by the British Heart Foundation (grant: FS/18/5/33319).

KDO was supported by UK Wellcome Trust Institutional Strategic Support Fund-LSHTM Fellowship 204928/Z/16/Z and Wellcome Trust-Royal Society Sir Henry Dale Fellowship 218554/Z/19/Z

AB is supported by research funding from NIHR, British Medical Association, Astra-Zeneca and UK Research and Innovation.

SD is supported by an Alan Turing Fellowship.

AGL is funded by the Wellcome Trust (204841/Z/16/Z), the National Institute for Health Research (NIHR) Great Ormond Street Hospital Biomedical Research Centre (19RX02) and the NIHR University College London Hospitals Biomedical Research Centre (BRC714a/HI/RW/101440).

RLB is an NIHR Research Professor.

HH is a NIHR Senior Investigator. His work is supported by: 1. Health Data Research UK (grant No. LOND1), which is funded by the UK Medical Research Council, Engineering and Physical Sciences Research Council, Economic and Social Research Council, Department of Health and Social Care (England), Chief Scientist Office of the Scottish Government Health and Social Care Directorates, Health and Social Care Research and Development Division (Welsh Government), Public Health Agency (Northern Ireland), British Heart Foundation and Wellcome Trust. 2. The BigData@Heart Consortium, funded by the Innovative Medicines Initiative-2 Joint Undertaking under grant agreement No. 116074 (also supporting AB and SD). This Joint Undertaking receives support from the European Union’s Horizon 2020 research and innovation programme and EFPIA; it is chaired, by DE Grobbee and SD Anker, partnering with 20 academic and industry partners and ESC. 3. The NIHR University College London Hospitals Biomedical Research Centre.

## References

1. Roberto CA, Swinburn B, Hawkes C, Huang TT, Costa SA, Ashe M, Zwicker L, Cawley JH, Brownell KD. Patchy progress on obesity prevention: emerging examples, entrenched barriers, and new thinking. Lancet. 2015;385(9985):2400–9

2. Lyn R, Heath E, Dubhashi J. Global Implementation of Obesity Prevention Policies: a Review of Progress, Politics, and the Path Forward. Curr Obes Rep. 2019 Dec;8(4):504–516

3. NCD-RisC, NCD Risk Factor Collaboration. Trends in adult body mass index in 200 countries from 1975 to 2014: a pooled analysis of 1698 population-based measurement studies with 19.2 million participants. 2016, Vol. 387, pp. 1377–96.

4. Ward ZJ, Bleich SN, Cradock AL, Barrett JL, Giles CM, Flax C, Long MW, Gortmaker SL. Projected U.S. State-Level Prevalence of Adult Obesity and Severe Obesity. N Engl J Med. 2019;381(25):2440–245.

5. NIHC, National Institute for Health and Care. Obesity in adults: prevention and lifestyle weight management programmes. [Online] 2016. https://www.nice.org.uk/guidance/qs111/resources/obesity-in-adults-prevention-and-lifestyle-weight-management-programmes-pdf-75545293071301 (retrieved 15/10/2019).

6. http://www.euro.who.int/data/assets/pdf_file/0003/247638/obesity-090514.pdf (retrieved 18/11/2020)

7. Dutton, G.R., Kim, Y., Jacobs, D.R., Jr., Li, X., Loria, C.M., Reis, J.P., Carnethon, M., Durant, N.H., Gordon_Larsen, P., Shikany, J.M., Sidney, S. and Lewis, C.E. (2016), 25□year weight gain in a racially balanced sample of U.S. adults: The CARDIA study. Obesity, 24: 1962–1968. doi:10.1002/oby.21573

8. Peter RS, Fromm E, Klenk J, Concin H, Nagel G. Change in height, weight, and body mass index: longitudinal data from Austria. Am J Hum Biol. 2014;26(5):690–696. doi:10.1002/ajhb.22582

9. Caman OK, Calling S, Midlöv P, Sundquist J, Sundquist K, Johansson SE. Longitudinal age-and cohort trends in body mass index in Sweden--a 24-year follow-up study. BMC Public Health. 2013;13:893. Published 2013 Sep 27. doi:10.1186/1471-2458-13-893

10. Bhaskaran K, Forbes HJ, Douglas I, et al. Representativeness and optimal use of body mass index (BMI) in the UK Clinical Practice Research Datalink (CPRD). BMJ Open 2013;3:e003389.

11. Nicholson, B.D., Aveyard, P., Bankhead, C.R. et al. Determinants and extent of weight recording in UK primary care: an analysis of 5 million adults’ electronic health records from 2000 to 2017. BMC Med. 2019;17(1):222.

12. Urgently needed: a framework convention for obesity control. Lancet. 2011 Aug 27;378(9793):741.

13. PHE, Public Health England. Adult obesity: applying All Our Health Public Health England 2015. [Online] (update 2019). https://www.gov.uk/government/publications/adult-obesity-applying-all-our-health/adult-obesity-applying-all-our-health (retrieved 6/12/2020).

14. Denaxas S, Gonzalez-Izquierdo A, Direk K, Fitzpatrick N, Fatemifar G, Banerjee A, Dobson R, Howe L, Kuan V, Lumbers T, Pasea L, Patel R, Shah A, Hingorani A, Sudlow C, and Hemingway H. UK phenomics platform for developing and validating electronic health record phenotypes: CALIBER. JAMIA. 2019;26(12):1545–59.

15. Fryar CD, Kruszon-Moran D, Gu Q, Ogden CL. Mean Body Weight, Height, Waist Circumference, and Body Mass Index Among Adults: United States, 1999-2000 Through 2015-2016. Natl Health Stat Report. 2018;(122):1–16.

16. Leacy FP, Floyd S, Yates TA. White IR.Analyses of Sensitivity to the Missing-at-Random Assumption Using Multiple Imputation With Delta Adjustment: Application to a Tuberculosis/HIV Prevalence Survey With Incomplete HIV-Status Data. Am J Epidemiol. 2017;185(4):304–15.

17. Fildes A, Charlton J, Rudisill C, Littlejohns P, Prevost AT, Gulliford MC. Probability of an Obese Person Attaining Normal Body Weight: Cohort Study Using Electronic Health Records. Am J Public Health. 2015 Sep;105(9):e54–9

18. Douglas IJ, Bhaskaran K, Batterham RL, Smeeth L. Bariatric Surgery in the United Kingdom: A Cohort Study of Weight Loss and Clinical Outcomes in Routine Clinical Care. PLoS Med. 2015 Dec 22;12(12):e1001925

19. Breland JY, Phibbs CS, Hoggatt KJ, Washington DL, Lee J, Haskell S, Uchendu US, Saechao FS, Zephyrin LC, Frayne SM. The Obesity Epidemic in the Veterans Health Administration: Prevalence Among Key Populations of Women and Men Veterans. J Gen Intern Med. 2017 Apr;32(Suppl 1):11–17.

20. Liu, Natalie MD∗; Birstler, Jen S†; Venkatesh, Manasa MS∗; Hanrahan, Lawrence P. PhD, MS‡; Chen, Guanhua PhD†; Funk, Luke M. MD, MPH∗;,© Weight Loss for Patients With Obesity, Medical Care: March 2020 - Volume 58 - Issue 3 - p 265–272

21. Hayba N, Partridge SR, Nour MM, Grech A, Allman Farinelli M. Effectiveness of lifestyle interventions for preventing harmful weight gain among young adults from lower socioeconomic status and ethnically diverse backgrounds: a systematic review. Obes Rev. 2018;19(3):333–346.

22. Averett SL, Sikora A, Argys LM. For better or worse: relationship status and body mass index. Econ Hum Biol. 2008 Dec;6(3):330–49. doi:10.1016/j.ehb.2008.07.003. Epub 2008 Jul 15. PMID: 18753018.

23. Vadeboncoeur C, Townsend N, Foster C. A meta-analysis of weight gain in first year university students: is freshman 15 a myth? BMC Obes. 2015 May 28;2:22

24. NIHC, National Institute for Health and Care. Obesity in adults: prevention and lifestyle weight management programmes. [Online] 2016. https://www.nice.org.uk/guidance/qs111/resources/obesity-in-adults-prevention-and-lifestyle-weight-management-programmes-pdf-75545293071301 (retrieved 15/10/2019).

25. Hayba N, Partridge SR, Nour MM, Grech A, Allman Farinelli M. Effectiveness of lifestyle interventions for preventing harmful weight gain among young adults from lower socioeconomic status and ethnically diverse backgrounds: a systematic review. Obes Rev. 2018;19(3):333–346.

26. Board on Children, Youth, and Families; Institute of Medicine; National Research Council. Improving the Health, Safety, and Well-Being of Young Adults: Workshop Summary. Washington (DC): National Academies Press (US); 2013 Sep 27. D, Background Paper: Pathways to Young Adulthood and Preventive Interventions Targeting Young Adults. Available from: https://www.ncbi.nlm.nih.gov/books/NBK202209/

27. Srivastav A.K., Sharma N., Samuel A.J. Impact of Coronavirus Disease-19 (COVID- 19) Lockdown on Physical Activity and Energy Expenditure Among Physiotherapy Professionals and Students Using Web-Based open E-Survey Sent through WhatsApp, Facebook and Instagram Messengers. Clin Epidemiol Glob Health. 2020 Jul 14 doi:10.1016/j.cegh.2020.07.003

28. Michail Katsoulis, Laura Pasea, Alvina Lai, Richard JB Dobson, Spiros Denaxas, Harry Hemingway, Amitava Banerjee. Obesity during the COVID-19 pandemic: cause of high risk or an effect of lockdown? A population-based electronic health record analysis in 1 958 184 individuals. Public Health (in press)

29. Simmonds M, Llewellyn A, Owen CG, Woolacott N. Predicting adult obesity from childhood obesity: a systematic review and meta-analysis. Obes Rev. 2016 Feb;17(2):95–107

30. Arvidsson D, Fridolfsson J, Börjesson M. Measurement of physical activity in clinical practice using accelerometers. J Intern Med. 2019;286(2):137–153.

