## Appendix for "Identifying high-risk groups for change in weight and body mass index: population cohort of 11 million measurements in 2.3 million adults"

**Contents**

**Section 1**

Evidence before this study from population-based cohorts of within person BMI change	page 2-4

**Section 2**

Additional results relevant for policy	page 5-16

**Section 3: Detailed statistical methods**

3.1: Calculation of the 1-, 5- and 10-year BMI change using window periods	page 17-20

3.2: Multiple imputation of BMI change under MNAR	page 21-27

3.3: Comparison of CPRD (electronic health records) versus Health Survey for England	page 28

3.4: Calculation of age-standardised transitions between BMI groups	page 29-32

3.5: Converting an odds ratio to a relative risk (for the online calculator)	page 33

**Section 4**

Sensitivity analysis	page 34-39

**Section 1: Evidence before this study from population-based cohorts of within person BMI change**

Table S1.1: Population-based cohort studies of within-individual BMI change in adults in relation to demographic factors. Cohorts with at least 5,000 participants and at least 1K young adults (18-24 years old), sorted by their sample size

| Author, publication year | Total N | N of young adults aged 18-24y† | BMI meas. after 2010 | Population based | EHR | Duration of follow up  1yr, 5yrs, 10yrs | Transitions between BMI categories  Normal weight to overweight / overweight to obese / obese to severe obese | Age range at baseline (years) | Additional demographic factors considered beyond age and sex? | |
| --- | --- | --- | --- | --- | --- | --- | --- | --- | --- | --- |
|  |  |  |  |  |  |  |  |  | Socioeconomic status | Ethnicity |
| **Our Study** | 2.3M | 200K | ● | ● | ● | ●●● | ●●● | 18-74 | ● | ● |
| Peter RS et al, 2014 [1] | 185K | 15K | ● | ● | ◌ | ◌●◌ | ◌◌◌ | 20-85 | ◌ | ◌ |
| Fildes et al. 2015 [2] | 166K | 10K | ● | ◌ | ● | ◌◌● | ◌◌◌ | ≥20 | ◌ | ◌ |
| Liu et al [3] | 60K | 8K | ● | ● | ◌ | ◌●◌ | ◌◌◌ | 18-70 | ◌ | ● |
| Droyvold et al, 2006 [4] | 45K | 1.5K | ◌ | ● | ◌ | ◌◌● | ◌◌◌ | ≥20 | ◌ | ◌ |
| Coogan PE et al, 2012 [5] | 23K | 5K | ◌ | ◌ | ◌ | ◌◌◌ | ◌◌◌ | 21-55 | ● | ● |
| Ouyang et al, 2015 [6] | 18K | 3K | ● | ● | ◌ | ◌●● | ◌◌◌ | 18-60 | ● | ◌ |
| Lebenbaum et al, 2018 [7] | 18K | 3K | ● | ● | ◌ | ◌◌● | ◌●◌ | ≥20 | ◌ | ● |
| Holowko et al [8] | 14K | 14K | ◌ | ◌ | ◌ | ◌◌◌ | ◌◌◌ | 18-23 | ● | ● |
| Paynter L. et al, 2015 [9] | 12K | 2K | ◌ | ● | ◌ | ◌●◌ | ◌◌◌ | 18-66 | ◌ | ◌ |
| Wilsgaard et al, 2005 [10] | 11K | 1K | ◌ | ◌ | ◌ | ◌●● | ◌◌◌ | 20-61 | ● | ◌ |
| Cllarke et al 2009 [11] | 11K | 1K | ◌ | ◌ | ◌ | ◌●● | ◌◌◌ | 18-45 | ● | ● |
| Malhatra et al 2013 [12] | 10K | 5K | ◌ | ◌ | ◌ | ◌●● | ◌◌◌ | 14-22 | ● | ● |
| Haheim et al, 2006 [13] | 7K | 1K | ◌ | ◌ | ◌ | ◌◌◌ | ◌◌◌ | 20-49 | ◌ | ◌ |
| Setia MS et al, 2009 [14] | 5.5K | 1K | ◌ | ● | ◌ | ●●● | ◌◌◌ | 18-54 | ◌ | ● |
| Caman OK et al, 2013 [15] | 5.5K | 1K | ◌ | ● | ◌ | ◌●● | ◌◌◌ | 16-71 | ◌ | ◌ |
| Lewis CE et al, 2000 [16] | 5.1K | 2.5K | ◌ | ◌ | ◌ | ◌◌● | ◌◌◌ | 18-32 | ◌ | ● |
| Dutton et al, 2016 [17] | 5K | 2.5K | ● | ◌ | ◌ | ◌●● | ◌◌◌ | 18-55 | ◌ | ● |

● feature present

◌ feature absent

†Estimation

**References**

1. Peter RS, Fromm E, Klenk J, Concin H, Nagel G. Change in height, weight, and body mass index: longitudinal data from Austria. *Am J Hum Biol*. 2014;26(5):690-696. doi:10.1002/ajhb.22582.
2. Fildes A, Charlton J, Rudisill C, Littlejohns P, Prevost AT, Gulliford MC. Probability of an Obese Person Attaining Normal Body Weight: Cohort Study Using Electronic Health Records. Am J Public Health. 2015 Sep;105(9):e54-9. doi: 10.2105/AJPH.2015.302773. Epub 2015 Jul 16. PMID: 26180980; PMCID: PMC4539812
3. Liu N, Birstler J, Venkatesh M, Hanrahan LP, Chen G, Funk LM. Weight Loss for Patients With Obesity: An Analysis of Long-Term Electronic Health Record Data. *Med Care*. 2020;58(3):265-272. doi:10.1097/MLR.0000000000001277
4. Drøyvold WB, Nilsen TI, Krüger O, et al. Change in height, weight and body mass index: Longitudinal data from the HUNT Study in Norway. *Int J Obes (Lond)*. 2006;30(6):935-939. doi:10.1038/sj.ijo.0803178
5. Coogan PE, Wise LA, Cozier YC, Palmer JR, Rosenberg L. Lifecourse educational status in relation to weight gain in African American women. *Ethn Dis*. 2012;22(2):198-206.
6. Ouyang Y, Wang H, Su C, et al. Use of quantile regression to investigate changes in the body mass index distribution of Chinese adults aged 18-60 years: a longitudinal study. BMC Public Health. 2015;15:278. Published 2015 Mar 21. doi:10.1186/s12889-015-1606-8
7. Lebenbaum M, Espin-Garcia O, Li Y, Rosella LC. Development and validation of a population based risk algorithm for obesity: The Obesity Population Risk Tool (OPoRT). PLoS One. 2018;13(1):e0191169
8. Holowko N, Jones M, Tooth L, Koupil I, Mishra G. Educational mobility and weight gain over 13 years in a longitudinal study of young women. *BMC Public Health*. 2014;14:1219. Published 2014 Nov 25. doi:10.1186/1471-2458-14-1219
9. Paynter L, Koehler E, Howard AG, Herring AH, Gordon-Larsen P. Characterizing long-term patterns of weight change in China using latent class trajectory modeling. *PLoS One*. 2015;10(2):e0116190
10. Wilsgaard T, Jacobsen BK, Arnesen E. Determining lifestyle correlates of body mass index using multilevel analyses: the Tromsø Study, 1979-2001. *Am J Epidemiol*. 2005;162(12):1179-1188. doi:10.1093/aje/kwi328
11. Clarke P, O'Malley PM, Johnston LD, Schulenberg JE. Social disparities in BMI trajectories across adulthood by gender, race/ethnicity and lifetime socio-economic position: 1986-2004. Int J Epidemiol. 2009;38(2):499-509. doi:10.1093/ije/dyn214
12. Malhotra R, Ostbye T, Riley CM, Finkelstein EA. Young adult weight trajectories through midlife by body mass category [published correction appears in Obesity (Silver Spring). 2014 Jul;22(7):1770]. *Obesity (Silver Spring)*. 2013;21(9):1923-1934. doi:10.1002/oby.20318
13. Lund Haheim L, Lund Larsen PG, Sogaard AJ, Holme I. Risk factors associated with body mass index increase in men at 28 years follow-up. *QJM*. 2006;99(10):665-671. doi:10.1093/qjmed/hcl090
14. Setia MS, Quesnel-Vallee A, Abrahamowicz M, Tousignant P, Lynch J. Convergence of body mass index of immigrants to the Canadian-born population: evidence from the National Population Health Survey (1994-2006). *Eur J Epidemiol*. 2009;24(10):611-623. doi:10.1007/s10654-009-9373-4
15. Caman OK, Calling S, Midlöv P, Sundquist J, Sundquist K, Johansson SE. Longitudinal age-and cohort trends in body mass index in Sweden--a 24-year follow-up study. BMC Public Health. 2013;13:893.
16. Lewis CE, Jacobs DR Jr, McCreath H, et al. Weight gain continues in the 1990s: 10-year trends in weight and overweight from the CARDIA study. Coronary Artery Risk Development in Young Adults. *Am J Epidemiol*. 2000;151(12):1172-1181. doi:10.1093/oxfordjournals.aje.a010167
17. Dutton, G.R., Kim, Y., Jacobs, D.R., Jr., Li, X., Loria, C.M., Reis, J.P., Carnethon, M., Durant, N.H., Gordon‐Larsen, P., Shikany, J.M., Sidney, S. and Lewis, C.E. (2016), 25‐year weight gain in a racially balanced sample of U .S . adults: The CARDIA study. Obesity, 24: 1962-1968. doi:[10.1002/oby.21573](https://doi.org/10.1002/oby.21573)

**Section 2: Additional results relevant for policy**

Figure S2.1A: Flow chart showing the number of individuals used in this study

**Individuals with linked EHR data and with at least 1 BMI & weight measurement in England from 1998 to 2016: N=2,396,540**

**Excluded individuals with implausible BMI measurements: N=68,093**

a) BMI recorded during pregnancy **N=64,909**

b) observations for which the maximum difference at the same date was more than 0.5 kg/m^2^ for BMI; **N=610**

c) individuals whose highest BMI was more than double compared to the BMI record, during 1998 and 2016. **N=321**

d) We excluded observations where the absolute between recorded and calculated BMI on the same date was more than 1 kg/m2. **N=2253**

Calculation of the BMI temporal trends and comparison with HSE (see Appendix)

**Individuals with at least 1 valid BMI measurement: N=2,328,477; N of BMI measurements: 11,187,383**

Figure S2.1B: Flow chart showing the number of individuals used for the calculation of 1-, 5- and 10-year BMI change

**Individuals with at least 1 valid BMI measurement: N=2,328,477; N of BMI measurements: 11,187,383**

**Excluded individuals aged ≥75yo: N=236,187**

**Individuals aged 18-74yo: N=2,092,260; N of BMI observations: 9,258,545**

**10-year** BMI change

**5-year** BMI change

**1-year** BMI change

Window period: **8 to 12 years**^†^

**Complete case analysis**

**N = 457,506**

Window period: **4 to 6 years**^†^

**Complete case analysis**

**N = 735,754**

Window period: **6 months to 2 years**^†^

**Complete case analysis**

**N = 1,001,872**

**Multiple imputation analysis^††^**

**N = 1,912,589 (^†††^**Excluded 179,671**)**

**Multiple imputation analysis^††^**

**N = 1,117,324 (^†††^**Excluded 974,936**)**

**Multiple imputation analysis^††^**

**N = 1,524,022 (^†††^**Excluded 411,287)

^†^ We selected all the pairs of BMI observations which were measured with time distance within the specified window periods. We then chose at random one pair of BMI observations, if an individual had >1 candidate pairs of BMI measurements for the calculation of BMI change

^††^For the rest individuals, we selected at random one time point with a BMI measurement and we applied multiple imputation

^†††^We excluded individuals who had follow-up time since last BMI measurement less than half of the period of interest (i.e. 6 months for the 1-year, 2.5 years for the 5-year and 5 years for the 10-year BMI change) or who died during the period of interest

Table S2.1: Baseline characteristics of individuals

|  | **Baseline characteristics for** | | |
| --- | --- | --- | --- |
|  | **1-year BMI change** | **5-year BMI change** | **10-year BMI change** |
| N of individuals | 1,912,589 | 1.524,022 | 1,117,324 |
| Age, in years; mean (sd) | 46.9 (15.5) | 47.2 (15.2) | 47.4 (14.8) |
| Women, % | 57.3% | 57.2% | 57.9% |
| BMI (in kg/m^2^); mean (sd)  Ethnicity  (missing: 20.2%, 19.6% 19.2% at 1, 5 and 10 years respectively)  White  Black  Asian  Mixed/Other    Index of multiple deprivation  (missing: 24.3%, 23.4%, 22.6% at 1, 5 and 10 years respectively)  1 (Less deprived)  2  3  4  5 (Most deprived)    Family history of CVD, % | 27.3 (5.9)  92.1%  4.0%  2.4%  1.5%  21.9%  21.5%  20.5%  19.1%  16.9%  25.3% | 27.3 (5.7)  93.0%  3.6%  2.1%  1.3%  22.4%  21.8%  20.5%  18.9%  16.5%  26.5% | 27.3 (5.6)  93.9%  3.3%  1.7%  1.1%  23.0%  22.0%  20.5%  18.7%  15.8%  28.0% |
| Use of Diuretics, % | 13.8% | 14.0% | 13.9% |
| **Prevalence of chronic diseases** |  |  |  |
| Cancer, % | 5.2% | 4.6% | 4.0% |
| CVD, % | 9.6% | 9.3% | 8.9% |
| Diabetes, % | 7.2% | 6.6% | 6.0% |
| Hypertension, % | 47.2% | 47.5% | 47.2% |
| Psychological disorders†, % | 30.3% | 28.5% | 26.8% |
| Other chronic diseases‡, % | 10.5% | 9.2% | 7.7% |

Figure S2.2: Distribution of the 1-, 5- and 10-year weight change* between 1998 and 2016 in England, by age group, separately in men (upper panel: with average height of 1.76m) and in women (lower panel: with height of 1.62m)

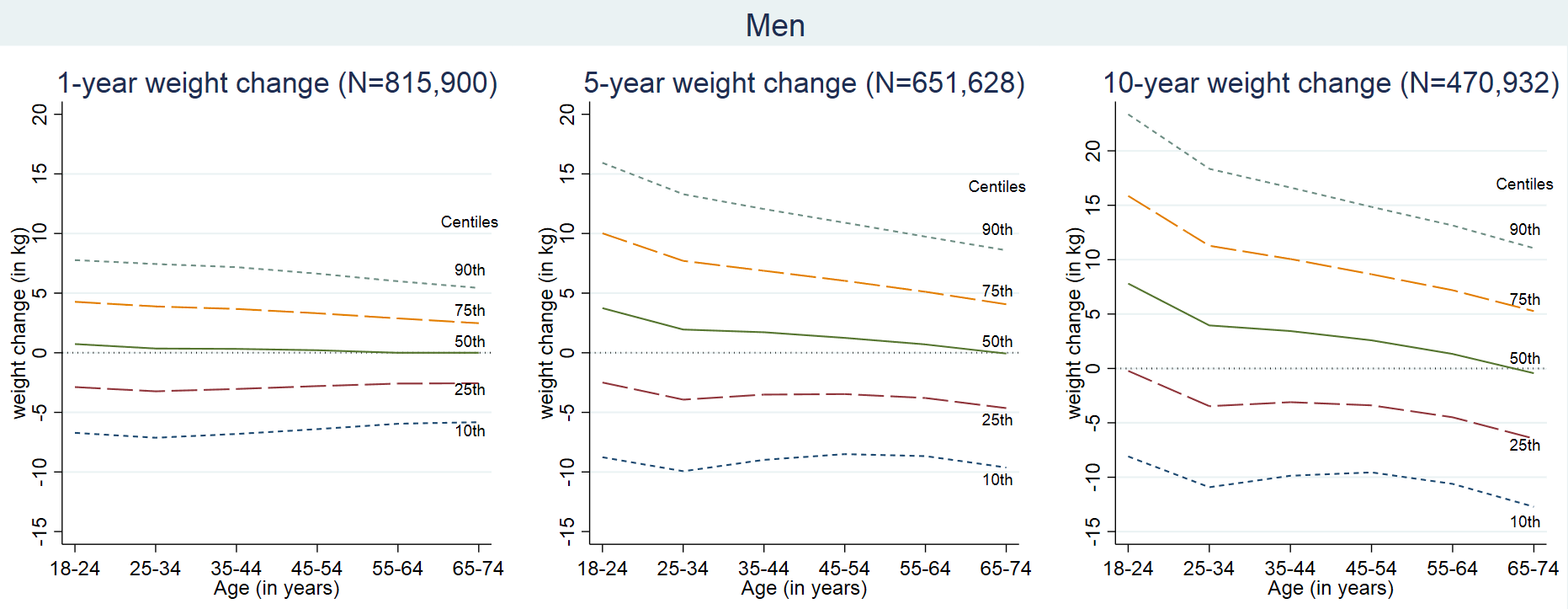

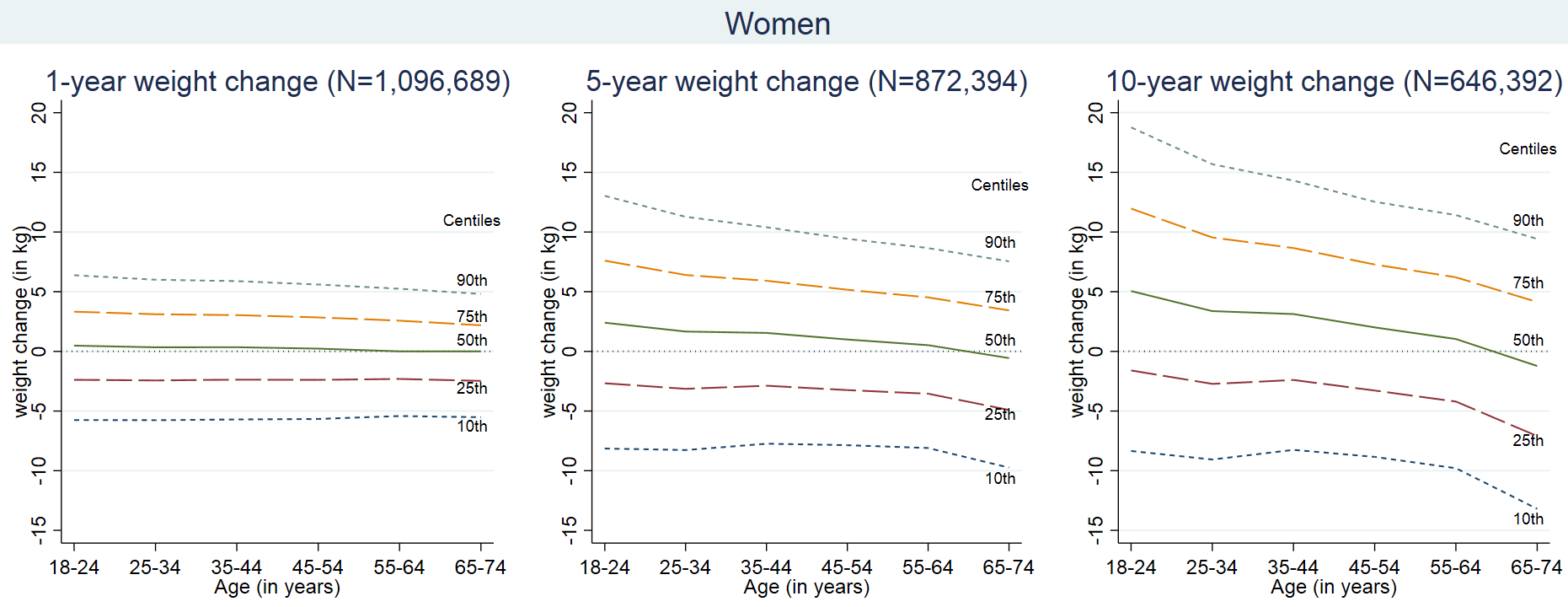

^*^Weight change is calculated indirectly from sex specific BMI change for men with height of 1.76m (median in CALIBER) and women with height of 1.62m (median in CALIBER)

Figure S2.3: Distributions of the 10-year weight change (in kg/m^2^) from 470,932 men (assuming average height of 1.76m) by age and BMI group

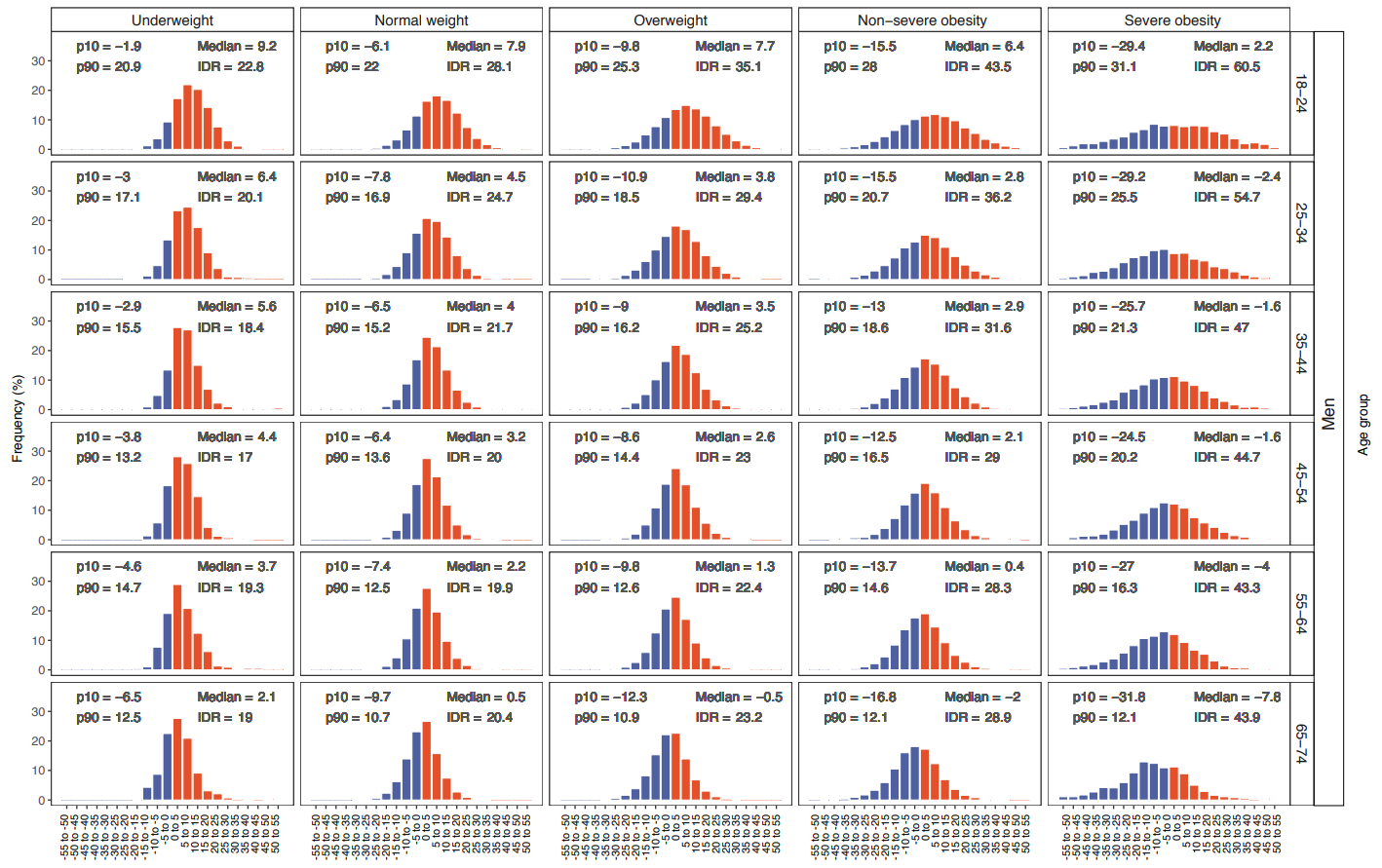

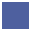
Weight loss
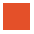
 Weight gain

Figure S2.4: Distributions of the 10-year weight change (in kg/m^2^) from 646,392 women (assuming average height of 1.62m) by age and BMI group

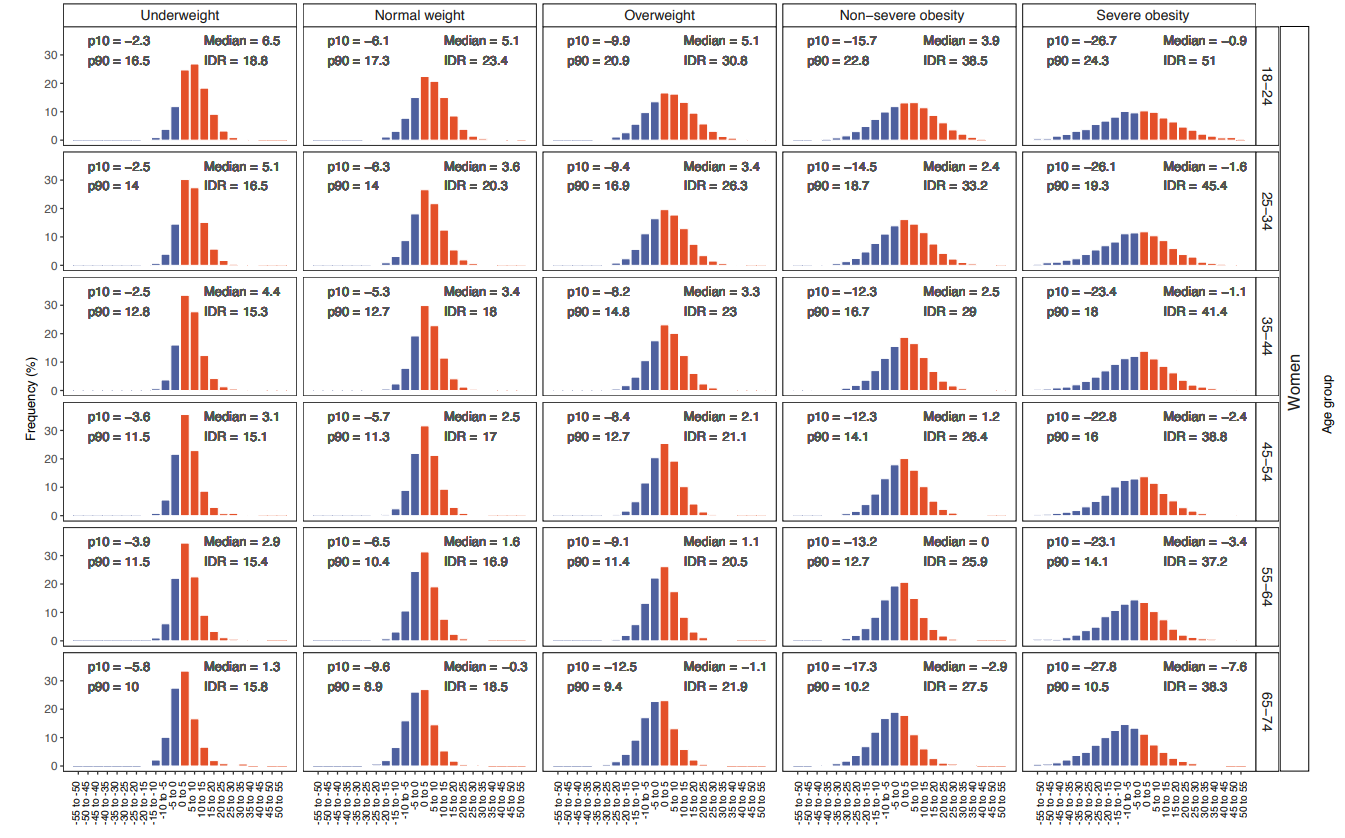

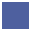
Weight loss
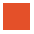
 Weight gain

Figure S2.5: Absolute risks and odds ratios* of transitioning at ten years from normal weight to overweight or obese, from overweight to obese and from non-morbid to morbid obese, by age, sex, ethnicity, social deprivation and region among young adults aged 18-24yo

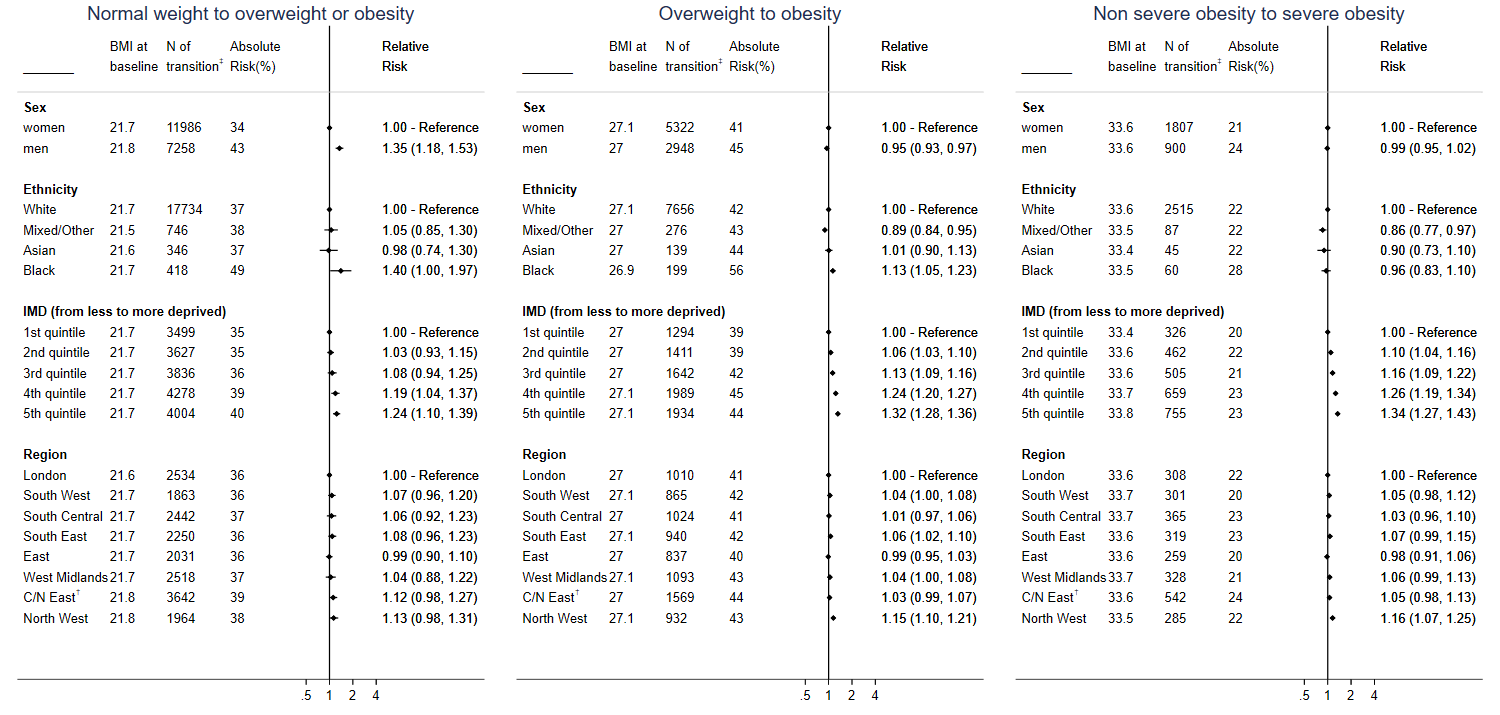

^*^Mutually adjusted for BMI (at baseline), gender, Index of multiple deprivation (IMD – quintiles; in categories), region, prevalence of mental disease (bipolar disorder, anxiety, acute stress, affective disorder, phobia, schizophrenia and depression), prevalence of chronic diseases (cancer, dementia, rheumatoid arthritis, diabetes, COPD, renal disease, gout, HIV, chronic kidney disease, inflammatory bowel disease, systemic lupus erythematosus and cardiovascular diseases), prevalence of hypertension, use of diuretics

†Central/North East

‡N of individuals who transitioned to higher BMI categories

Figure S2.6: Absolute risks and odds ratios* of remaining obese at ten years, by age, sex, ethnicity, social deprivation and region

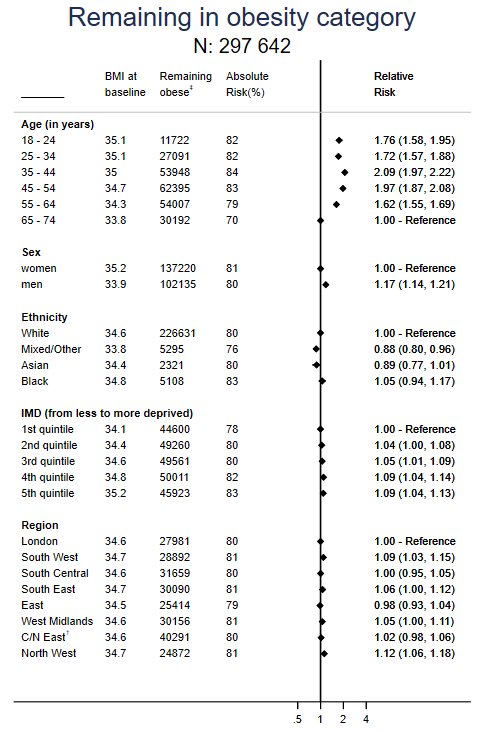

^*^Mutually adjusted for BMI (at baseline), age group, gender, Index of multiple deprivation (IMD – quintiles; in categories), region, prevalence of mental disease (bipolar disorder, anxiety, acute stress, affective disorder, phobia, schizophrenia and depression), prevalence of chronic diseases (cancer, dementia, rheumatoid arthritis, diabetes, COPD, renal disease, gout, HIV, chronic kidney disease, inflammatory bowel disease, systemic lupus erythematosus and cardiovascular diseases), prevalence of hypertension, use of diuretics

†Central/North East

‡N of individuals who remained obese

Figure S2.7: Absolute risks and odds ratios* of transitioning at one year from normal weight to overweight or obese, from overweight to obese and from non-morbid to morbid obese, by age, sex, ethnicity, social deprivation and region

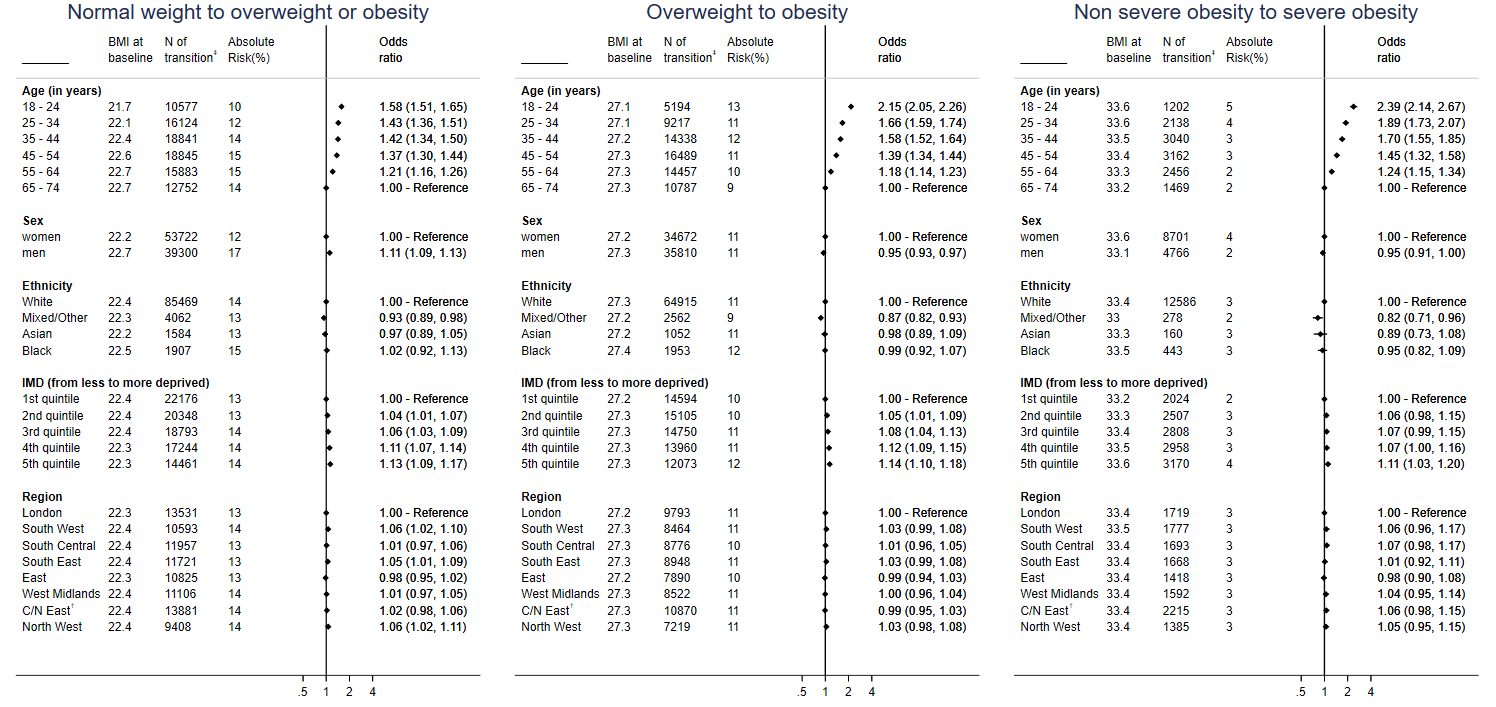

^*^Mutually adjusted for BMI (at baseline), age group, gender, Index of multiple deprivation (IMD – quintiles; in categories), region, prevalence of mental disease (bipolar disorder, anxiety, acute stress, affective disorder, phobia, schizophrenia and depression), prevalence of chronic diseases (cancer, dementia, rheumatoid arthritis, diabetes, COPD, renal disease, gout, HIV, chronic kidney disease, inflammatory bowel disease, systemic lupus erythematosus and cardiovascular diseases), prevalence of hypertension, use of diuretics

†Central/North East

‡N of individuals who transitioned to higher BMI categories

Figure S2.8: Absolute risks and odds ratios* of transitioning at five years from normal weight to overweight or obese, from overweight to obese and from non-morbid to morbid obese, by age, sex, ethnicity, social deprivation and region

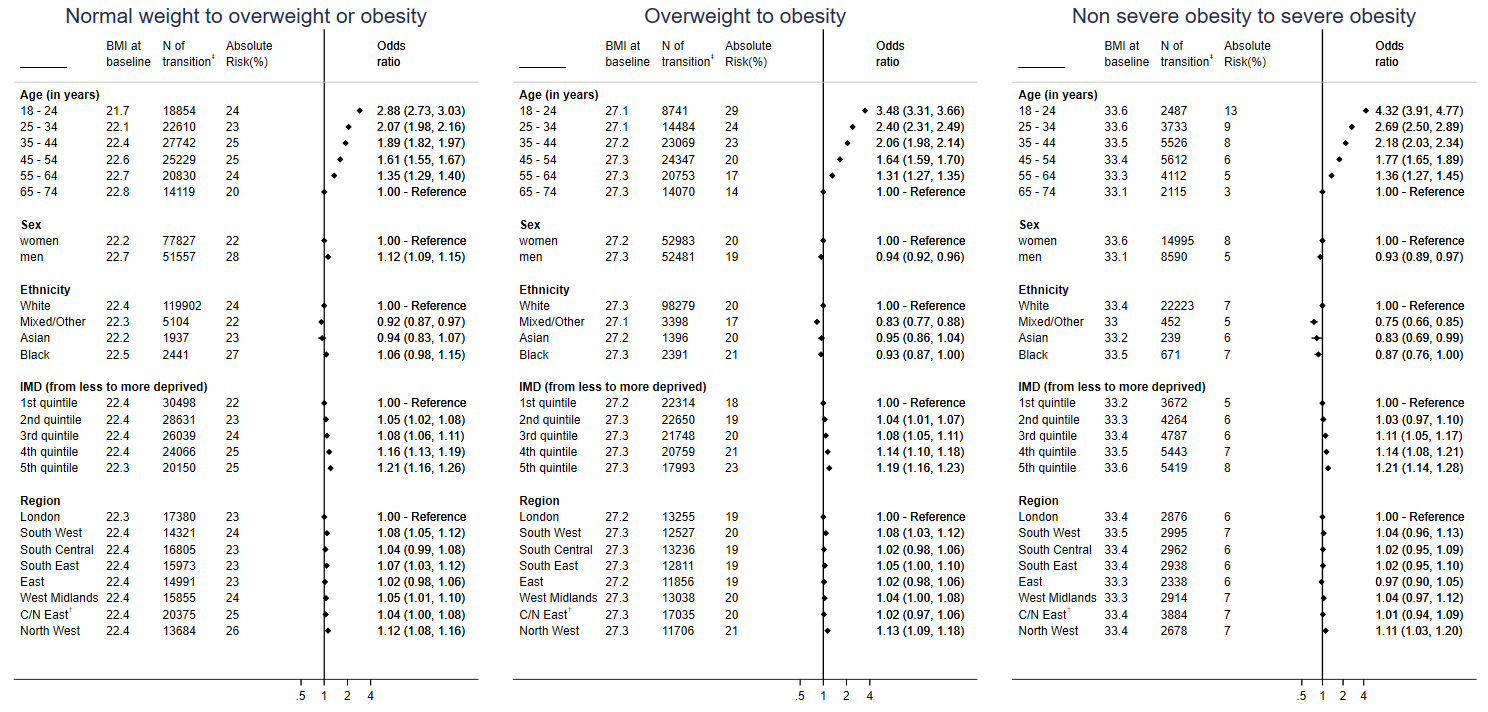

^*^Mutually adjusted for BMI (at baseline), age group, gender, Index of multiple deprivation (IMD – quintiles; in categories), region, prevalence of mental disease (bipolar disorder, anxiety, acute stress, affective disorder, phobia, schizophrenia and depression), prevalence of chronic diseases (cancer, dementia, rheumatoid arthritis, diabetes, COPD, renal disease, gout, HIV, chronic kidney disease, inflammatory bowel disease, systemic lupus erythematosus and cardiovascular diseases), prevalence of hypertension, use of diuretics

†Central/North East

‡N of individuals who transitioned to higher BMI categories

Figure S2.9: Absolute risk for the transitions from normal weight to overweight or obese, from overweight to obese and from non-morbid to morbid obese in 1 year, by age, social deprivation, sex and initial BMI

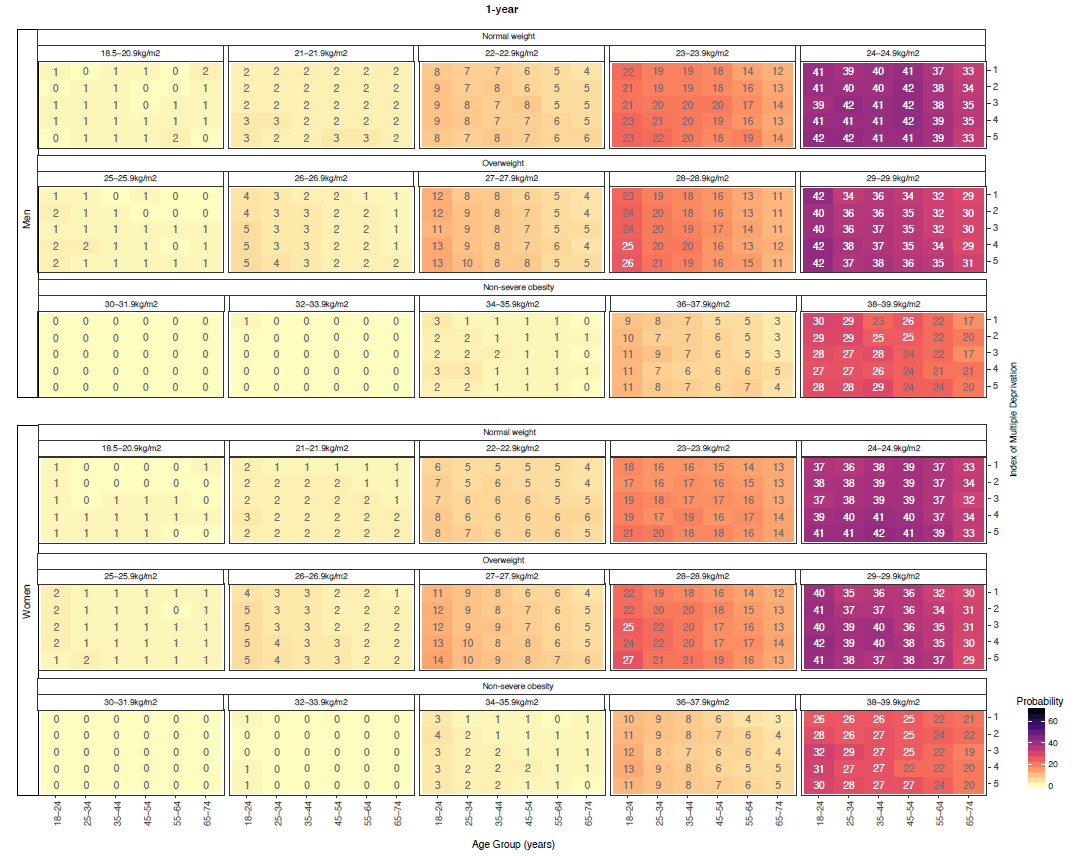

Figure S2.10: Absolute risk for the transitions from normal weight to overweight or obese, from overweight to obese and from non-morbid to morbid obese in 5 years, by age, social deprivation, sex and initial BMI

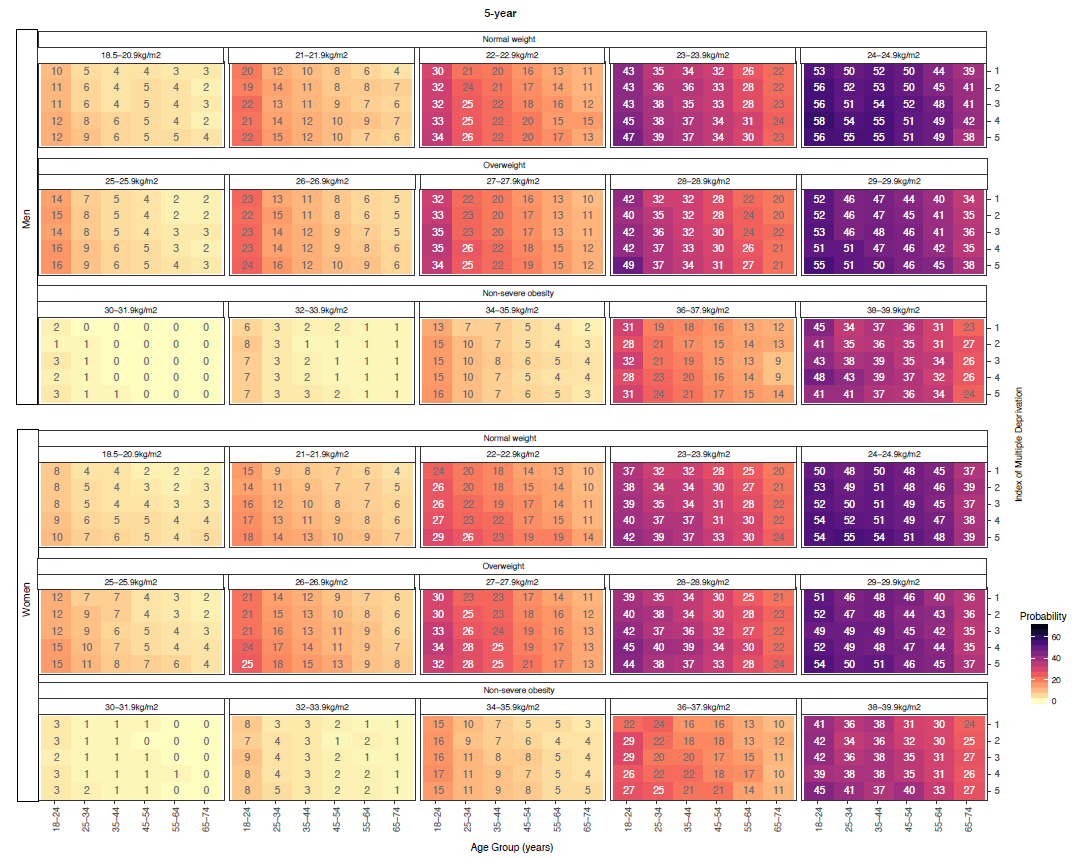

**Section 3: Detailed statistical methods**

**3.1. Calculation of the 1-, 5- and 10-year BMI change using window periods**

To estimate the 1-, 5- and 10-year BMI change periods, we used window periods (6 months-2 years, 4 years-6 years and 8 years-12 years respectively) in which individuals had at least 2 BMI measurements (see table S1.1).

**Table S3.1.1: Window periods used to select a pair of BMI observations and estimate BMI change**

| **Estimated Period** | **Window period** |
| --- | --- |
| 1-year BMI change | 6 months to 2 years |
| 5-year BMI change | 4 years to 6 years |
| 10-year BMI change | 8 years to 12 years |

If an individual had more than one pair of BMI measurements, we selected at random one of them. To estimate the 1-, 5- and 10-year BMI change, we assume that BMI change is linear in this window, i.e. if an individual’s BMI decreased by 2.6 kg/m^2^ in 1.3 years, we assume that the 1-year BMI change for this individual in -2 kg/m^2^.

Below, we present an example of a hypothetical individual, her BMI measurements between 1998 and 2016 and we explain how her BMI measurements contribute in the calculation of the 1-, 5- and 10-year BMI change. Her BMI was measured at 27.2 kg/m^2^ in 23/11/2001, at 27.2 kg/m^2^ in 8/10/2002, at 27.2 kg/m^2^ in 2/4/2003 and at 27.2 kg/m^2^ in 28/3/2009 (see below in Figure S1.1).

**Figure S3.1.1: Estimating BMI change for a hypothetic individual**

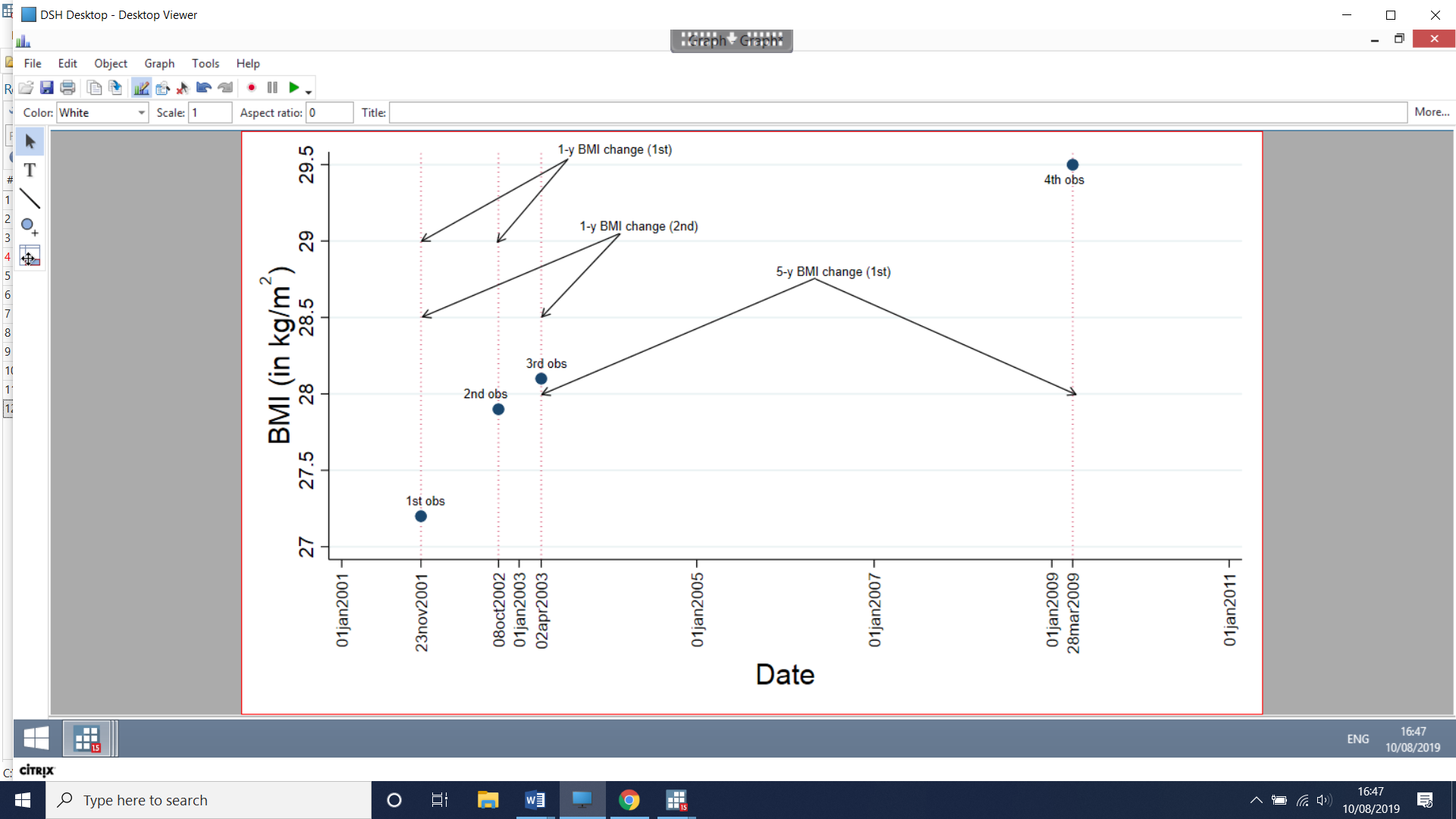

From her data, the pairs of BMI observation that contribute to the 1-year BMI change (i.e. within 6 months to 2 years) are

1. (1^st^, 2^nd^) BMI measurements
2. (1^st^, 3^rd^) BMI measurements

The pairs of BMI observation that contribute to the 5-year BMI change (i.e. within 4 to 6 years) are

1. (3^rd^, 4^th^) BMI measurements

Additionally, she has no pairs of BMI measurements which contribute to the 5-year BMI change (i.e. within 8 to 12 years) are

To estimate her contribution to the 1-, 5- and 10-year BMI change, we select at random one pair of BMI observations for each period. So, for example, we select at random her 2^nd^ pair of BMI measurements, i.e. (1^st^, 3^rd^) observations. For the calculation of the 5-year BMI change, we have only 1 pair of BMI measurements, i.e. (3^rd^, 4^th^) observations. Moreover, we assumed linear trend within each period, so her contribution to the 1-year BMI change is 0.18% and to the 5-year BMI change is 0.32% (see table S1.2). Finally, she will have a missing value for the calculation of the 10-year BMI change.

**Table S3.1.2: Estimation of BMI change of a hypothetical individual (from figure S1.1)**

| **Estimated Period** | **Pairs of BMI observations contributing** | **Pair selected** | **Period between the two BMI measurements** | **Observed BMI change** | **Estimated BMI change** |
| --- | --- | --- | --- | --- | --- |
| 1-year BMI change | (1^st^, 2^nd^), (1^st^, 3^rd^) | (1^st^, 3^rd^) | 1.36 years | 0.25% | 0.25% / 1.36 = **0.18%** |
| 5-year BMI change | (3^rd^, 4^th^) | (3^rd^, 4^th^) | 5.98 years | 0.38% | 5* (0.38% /5.98) = **0.32%** |
| 10-year BMI change | - | - | - | - | - |

**3.2 Multiple imputation of BMI change under MNAR**

In the analysis of 1-, 5-, 10-year BMI change, there were many individuals that had no pair of BMI measurements within the corresponding window period, so these individuals had a missing value for BMI change. For example, the hypothetical individual in figure S1.1, she had no pair of BMI measurements between 8 and 12 years, thus resulting without an estimate for the 10-year BMI change.

To tackle the problem of missing values in our analysis, we assumed that the missingness mechanism for BMI change was missing not at random (MNAR). In other words, missing values on BMI change might depend on BMI change itself, as it is more likely that those individuals who don’t weigh in frequently in their practice, are healthy and their weight/BMI has not changed. For this reason, we applied multiple imputation with delta adjustment, which is a 2-part method: in the 1^st^ part, we apply multiple imputation for BMI change (and for other covariates), as if missing at random (MAR) held, but then, in the 2^nd^ part, we added delta values to our imputed datasets of BMI change. To account for the fact that some individuals had multiple contacts with the health system, and multiple measurements, we considered a summary variable that recorded the number of visits recorded. More specifically,

1. We created 10 copies of datasets in which missing values of BMI change, ethnicity, index of multiple deprivation (IMD), smoking status and physical activity were replaced by imputed values sampled from their predictive distribution, through multiple imputation by chained equations, which was applied separately in 6 age groups (18-24 years old, 25-34 years old, 35-44 years old, 45-54 years old, 55-64 years old and 65-74 years old). The imputation model in each age group included
   a) Use of diuretics, history of bariatric surgery, prevalence of hypertension, cancer (apart from non-melanoma skin cancer), cardiovascular disease, diabetes, other chronic diseases (dementia, chronic kidney disease, systemic lupus erythematosus, rheumatoid arthritis, gout, ulcerative colitis, Parkinson disease, multiple sclerosis, renal disease, renal failure) and psychological conditions (depression, anxiety, stress, phobia, schizophrenia, bipolar disorder, affective disorder).
   b) For each of the 1-, 5- and 10-year period, we additionally considered the use of diuretics and the performance of bariatric surgery during these periods, as well as the presence of all the incident chronic conditions described in a) above,
   c) We included ethnicity, index of multiple deprivation, physical activity and smoking status that also had missing values. We considered physical activity and smoking status after 1 or 5 or 10 years after the BMI measurement, depending on the imputation of the corresponding BMI change
   d) We also included age, quadratic age, cubic age, time (from 1998 to year of baseline), quadratic time (from 1998 to year of baseline), family history of CVD
   e) BMI at baseline in each BMI category [5 coefficients were created, each for underweight, normal weight, overweight, obese (BMI≥30 & BMI<40kg/m^2^) and morbid obese (BMI≥40 kg/m^2^)]
   f) We added
      i. one variable for the frequency of contacts of an individual with the health care system during the corresponding period (i.e. one, five and ten years). More specifically, this variable had 6 categories and was modelled as ordered (1: 1 time, 2: 2-3 times, 3: 4-5 times, 4: 6-11 times, 5: 12-23 times and 6: 24 or more times)
      ii. one variable for the frequency of weight measurements in the primary care during the corresponding period (i.e. one, five and ten years). More specifically, this variable had 6 categories and was modelled as ordered (1: 1 time, 2: 2 times, 3: 3 or more times)
2. We then added delta values to our imputed datasets of BMI change, to address the problem of the missing not at random mechanism. The delta values were derived as follows:

We used the estimations of BMI levels from both men and women from the Health Survey from England between 1998 and 2016 for the following age groups: 16-24, 25-34, 35-44, 45-54, 55-64, 65-74 and 75+ years old. We then calculated the 10-year BMI change for each age group, for each year separately. For example, the average 10-year BMI change in men aged 25-34 in 1998, noted $\bar{BMI\_ch25\_34}$, was

$$\bar{BMI\_ch25\_34(1998)}=\frac{\left( \bar{BMI25\_34(2008)}-\bar{BMI25\_34(1998)} \right)}{\bar{BMI25\_34(1998)}}=\frac{27.8-26.1}{26.1}=6.5\%$$

In table S2.1, we show which BMI estimates we combine to calculate the 10-year BMI changes for men aged 25-34, from HSE, for all the years from 1998 to 2006.

**Table S3.2.1: BMI levels of men estimated from the Health Survey for England (HSE).**

| **Year** | **Age**  **16-24** | **Age**  **25-34** | **Age**  **35-44** | **Age**  **45-54** | **Age**  **55-64** | **Age**  **65-74** | **Age**  **75+** |
| --- | --- | --- | --- | --- | --- | --- | --- |
| **1998** | 23.5 | 26.1 | 26.7 | 27.4 | 27.8 | 27.5 | 26.4 |
| **1999** | 23.2 | 25.9 | 27.0 | 27.4 | 27.3 | 27.2 | 26.6 |
| **2000** | 23.4 | 26.7 | 27.4 | 27.5 | 27.9 | 27.7 | 26.5 |
| **2001** | 24.1 | 26.4 | 27.4 | 27.9 | 27.9 | 27.7 | 26.7 |
| **2002** | 23.8 | 26.6 | 27.3 | 27.9 | 27.9 | 28.1 | 27.1 |
| **2003** | 23.7 | 26.3 | 27.6 | 28.0 | 28.0 | 28.1 | 27.1 |
| **2004** | 23.9 | 26.3 | 27.8 | 28.2 | 28.3 | 28.0 | 26.9 |
| **2005** | 23.7 | 26.5 | 27.9 | 28.0 | 28.1 | 27.9 | 26.8 |
| **2006** | 24.1 | 26.7 | 27.8 | 28.0 | 28.6 | 28.3 | 27.0 |
| **2007** | 24.1 | 26.1 | 27.7 | 28.6 | 28.4 | 28.2 | 27.1 |
| **2008** | 23.7 | 26.5 | 27.8 | 28.1 | 28.5 | 28.7 | 27.4 |
| **2009** | 23.8 | 25.7 | 27.2 | 28.7 | 28.6 | 28.4 | 27.2 |
| **2010** | 24.4 | 26.3 | 28.1 | 28.8 | 28.9 | 28.3 | 27.8 |
| **2011** | 24.0 | 26.2 | 27.1 | 28.6 | 28.6 | 28.5 | 28.1 |
| **2012** | 24.4 | 26.0 | 27.7 | 28.8 | 28.6 | 28.5 | 27.7 |
| **2013** | 23.7 | 26.7 | 28.0 | 28.8 | 28.6 | 28.6 | 27.6 |
| **2014** | 23.8 | 26.4 | 27.7 | 28.4 | 28.9 | 27.9 | 28.0 |
| **2015** | 25.2 | 26.5 | 27.8 | 28.6 | 29.1 | 28.7 | 27.5 |
| **2016** | 24.0 | 26.4 | 27.9 | 28.7 | 28.8 | 28.6 | 27.6 |

*From the circles connected by an arrow, we can derive the average 10-year BMI change for men aged 25-34, for all the years from 1998-2006*

**Table S3.2.2: BMI levels of women estimated from the Health Survey for England (HSE).**

| **Year** | **Age**  **16-24** | **Age**  **25-34** | **Age**  **35-44** | **Age**  **45-54** | **Age**  **55-64** | **Age**  **65-74** | **Age**  **75+** |
| --- | --- | --- | --- | --- | --- | --- | --- |
| **1998** | 23.8 | 25.5 | 26.4 | 27.0 | 27.6 | 27.8 | 26.4 |
| **1999** | 23.7 | 25.4 | 26.2 | 27.1 | 27.8 | 27.9 | 26.6 |
| **2000** | 23.6 | 25.6 | 26.4 | 27.2 | 28.3 | 28.1 | 26.9 |
| **2001** | 24.1 | 25.8 | 26.6 | 27.6 | 28.1 | 27.9 | 26.7 |
| **2002** | 24.1 | 26.0 | 26.9 | 27.5 | 27.8 | 27.7 | 27.0 |
| **2003** | 24.2 | 26.0 | 26.7 | 27.4 | 27.8 | 28.1 | 27.3 |
| **2004** | 24.4 | 25.7 | 26.8 | 27.4 | 28.2 | 28.0 | 26.9 |
| **2005** | 24.3 | 25.8 | 27.0 | 27.8 | 27.8 | 28.2 | 27.1 |
| **2006** | 24.0 | 25.9 | 26.8 | 27.6 | 28.0 | 28.6 | 27.5 |
| **2007** | 24.0 | 25.8 | 27.0 | 27.4 | 28.0 | 28.0 | 27.2 |
| **2008** | 24.3 | 25.8 | 27.1 | 27.7 | 28.0 | 28.5 | 27.2 |
| **2009** | 24.8 | 25.9 | 27.2 | 27.6 | 28.2 | 28.5 | 27.2 |
| **2010** | 24.2 | 26.2 | 27.2 | 28.0 | 28.4 | 29.0 | 27.4 |
| **2011** | 24.2 | 26.2 | 27.4 | 28.0 | 28.1 | 28.2 | 28.0 |
| **2012** | 24.6 | 26.3 | 27.0 | 27.8 | 28.1 | 28.1 | 27.5 |
| **2013** | 24.3 | 26.0 | 27.3 | 27.6 | 28.2 | 28.2 | 27.2 |
| **2014** | 24.4 | 26.2 | 27.2 | 28.1 | 28.6 | 28.4 | 27.6 |
| **2015** | 24.8 | 26.4 | 27.0 | 28.0 | 28.4 | 28.0 | 27.6 |
| **2016** | 24.5 | 25.9 | 27.1 | 28.4 | 28.3 | 28.4 | 27.8 |

We then calculated the average of all the BMI changes between 1998 and 2006 for all age groups, in men and women separately. Of note, we could not calculate BMI change for those individuals aged ≥75 years old. The values for average 10-year BMI change values are presented below, in table S2.3.

**Table S3.2.3: Average 10-year BMI change (%) values for both men and women, estimated from the Health Survey for England (HSE).**

| **Sex** | **Age**  **16-24** | **Age**  **25-34** | **Age**  **35-44** | **Age**  **45-54** | **Age**  **55-64** | **Age**  **65-74** |
| --- | --- | --- | --- | --- | --- | --- |
| **Men** | 10.9% | 5.0% | 4.2% | 3.2% | 1.7% | -0.1% |
| **Women** | 8.6% | 5.4% | 4.8% | 3.1% | 1.6% | -0.2% |

Moreover, we consider all the average 10-year BMI change calculated by the HSE as the reference BMI changes that should match with the ones from CALIBER. For this reason, we name the following variables:

***BMI_ch_ref_1_*** *🡪 Average BMI change in men aged between 16-24yo*

***R_1_****🡪 Observed values of BMI change in men aged between 16-24yo in CALIBER*

***BMI_ch_ref_2_*** *🡪 Average BMI change in men aged between 25-34yo*

***R_2_****🡪 Observed values of BMI change in men aged between 25-34yo in CALIBER*

…

***BMI_ch_ref_6_*** *🡪 Average BMI change in men aged between 65-74yo*

***R_6_****🡪 Observed values of BMI change in men aged between 65-74yo in CALIBER*

and we continue with the women, i.e.

***BMI_ch_ref_7_*** *🡪 Average BMI change in women aged between 16-24yo*

***R_7_****🡪 Observed values of BMI change in women aged between 16-24yo in CALIBER*

***BMI_ch_ref_8_*** *🡪 Average BMI change in women aged between 25-34yo*

***R_8_****🡪 Observed values of BMI change in women aged between 25-34yo in CALIBER*

…

***BMI_ch_ref_12_*** *🡪 Average BMI change in women aged between 65-74yo*

***R_12_****🡪 Observed values of BMI change in women aged between 65-74yo in CALIBER*

We also set **BMI_ch_1_ – BMI_ch_12_** the estimated BMI change per age group and sex, **n_obs_1_ - n_obs_12_** the number of individuals with observed BMI change values per age group and sex in CALIBER, as well as **n_mis_1_ - n_mis_12_** the number of individuals with missing values on BMI change per age group and sex in CALIBER.

We then require that the average 10-year BMI estimates per age group and age from CALIBER, after multiple imputation, would be the same with the corresponding estimates from HSE. In other words, for each i=1,2,…,12, we set

$\frac{{n\_obs}_{i}*\left( \bar{{BMI\_ch}_{i}*R_{i}} \right)+{n\_mis}_{i}*\left( \bar{{BMI\_ch}_{i}*\left( 1-R_{i} \right)}*+\delta_{i} \right)}{{n\_obs}_{i}+{n\_mis}_{i}}={BMI\_ch\_ref}_{i}$	(1)

If we solve the above equation (1) for $\delta_{i}$, we have that

$\delta_{i}=\frac{{BMI\_ch\_ref}_{i}*\left( {n\_obs}_{i}+{n\_mis}_{i} \right)-{n\_obs}_{i}*\left( \bar{{BMI\_ch}_{i}*R_{i}} \right)}{{n\_mis}_{i}}-\bar{{BMI\_ch}_{i}*\left( 1-R_{i} \right)}$	(2)

The delta values for the average 10-year BMI change are presented in table S2.4 below

**Table S3.2.4: δ-values for the average 10-year BMI change values**

| **Sex** | **Age**  **16-24** | **Age**  **25-34** | **Age**  **35-44** | **Age**  **45-54** | **Age**  **55-64** | **Age**  **65-74** |
| --- | --- | --- | --- | --- | --- | --- |
| **Men** | -3,3% | -2,5% | -0.3% | 0.7% | 0,0% | 0.6% |
| **Women** | -4,2% | -4,3% | -2.4% | -1.3% | -0.3% | -1.0% |

To calculate the delta values for the 1-year and 5-year BMI change, we worked as follows. For the 1-year BMI change, we estimated the average 1-year BMI change from HSE, by assuming that

$\left( 1+ {\mathrm{BMI}_{\mathrm{ch}_{\mathrm{ref}}}}_{i}^{10year}(\%) \right)=\left( 1+ {BMI\_ch\_ref}_{i}^{1year}(\%) \right)^{10}$, i.e

$\left( 1+ {\mathrm{BMI}_{\mathrm{ch}_{\mathrm{ref}}}}_{i}^{1year}(\%) \right)=\sqrt[10]{\left( 1+ {BMI\_ch\_ref}_{i}^{10year}(\%) \right)}$

So, the delta values for the 1-year BMI change are presented in table S2.5 below

**Table S3.2.5: δ-values for the average 1-year BMI change values**

| **Sex** | **Age**  **16-24** | **Age**  **25-34** | **Age**  **35-44** | **Age**  **45-54** | **Age**  **55-64** | **Age**  **65-74** |
| --- | --- | --- | --- | --- | --- | --- |
| **Men** | -0.8% | -0.6% | -0.1% | 0.0% | 0.0% | 0.2% |
| **Women** | -1.8% | -0.8% | -0.7% | -0.5% | 0.1% | -0.1% |

and

($1+ {BMI\_ch\_ref}_{i}^{10year}(\%)$)=$\left( 1+ {BMI\_ch\_ref}_{i}^{5year}(\%) \right)^{2}$, i.e

($1+ {BMI\_ch\_ref}_{i}^{5year}(\%)$)=$\sqrt{\left( 1+ {BMI\_ch\_ref}_{i}^{10year}(\%) \right)}$

So, the delta values for the 5-year BMI change are presented in table S2.6 below

**Table S3.2.6: δ-values for the average 5-year BMI change values**

| **Sex** | **Age**  **16-24** | **Age**  **25-34** | **Age**  **35-44** | **Age**  **45-54** | **Age**  **55-64** | **Age**  **65-74** |
| --- | --- | --- | --- | --- | --- | --- |
| **Men** | -2.2% | -1.6% | -0.2% | 0.1% | -0.5% | -0.1% |
| **Women** | -3.4% | -2.6% | -2.1% | -1.8% | -0.9% | -1.7% |

**3.3: Comparison of CPRD (electronic health records) versus Health Survey for England**

Figure S3.3.1: Estimates of mean BMI levels in England between 1998 and 2016 from Electronic Health Records (EHR) and Health Survey from England (HSE), by age and sex

Figure might change

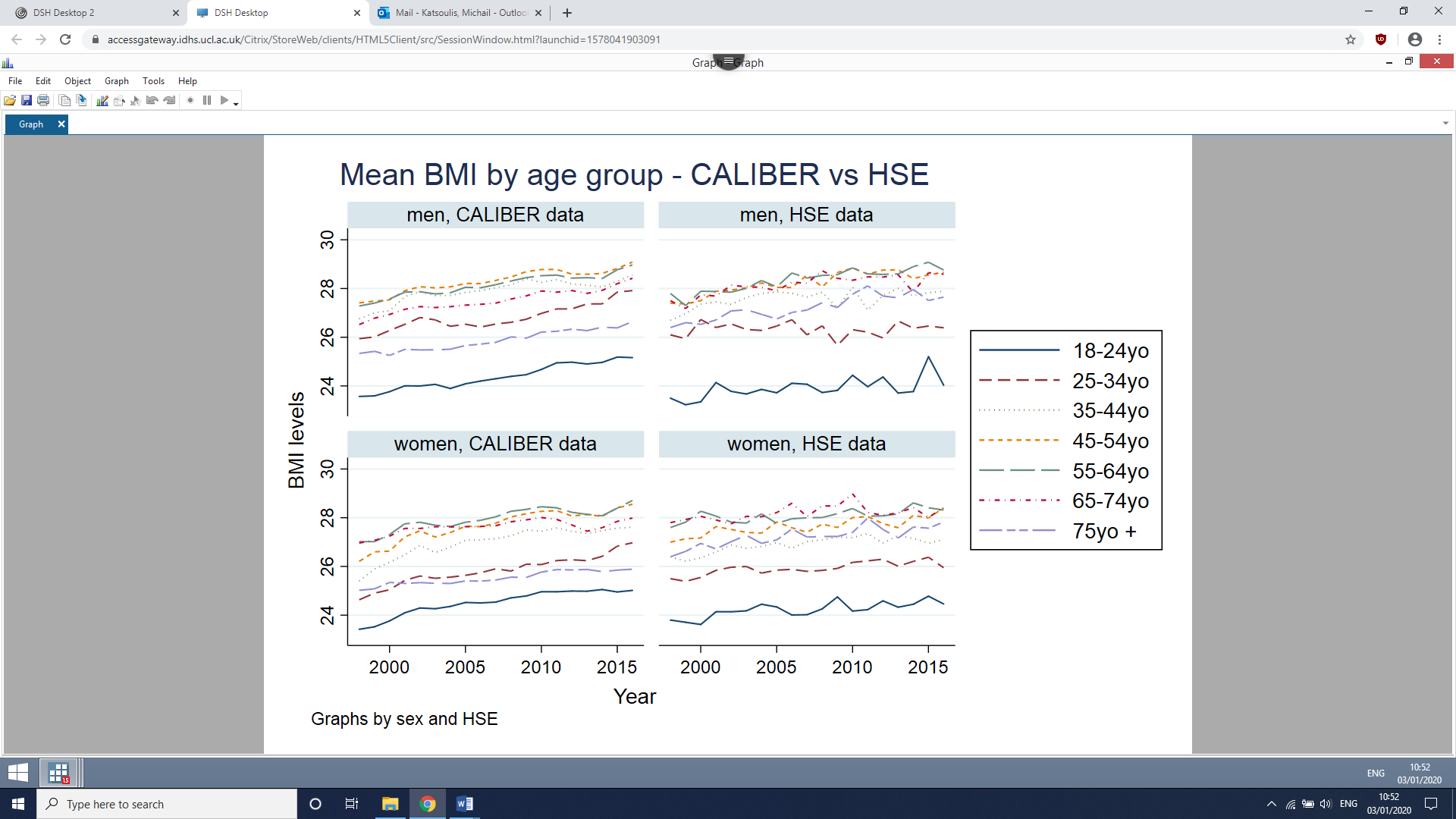

**3.4: Calculation of age-standardised transitions between BMI groups**

We calculated the age-standardised transitions between normal weight, overweight and obesity, after taking into consideration the age structure of the English population from ONS between 1998 and 2016. Specifically, we calculated two different weights, which correspond to: i. the proportion of each age group among individuals aged 18-74 (see table S6.1) and ii. the prevalence of normal weight, overweight and obesity for each age group (see table S6.2)

*Table S3.4.1: Age structure^†^ of the English population from ONS between 1998 and 2016*

| **Age group**  **(in years)** | **Proportion among individuals aged 18-74yo** |
| --- | --- |
| 18-24 | 12.7% |
| 25-34 | 19.7% |
| 35-44 | 20.4% |
| 45-54 | 19.0% |
| 55-64 | 16.0% |
| 65-74 | 12.3% |

^†^ Average proportion of each age group among individuals aged 18-74, between 1998 and 2016

*Table S3.4.2: BMI structure^†^ of the English population from HSE between 1998 and 2016, by age group*

| **Age group**  **(in years)** | **Proportion of normal weight** | **Proportion of overweight** | **Proportion of obesity** |
| --- | --- | --- | --- |
| 18-24 | 60.8% | 22.2% | 10.7% |
| 25-34 | 46.4% | 33.5% | 18.3% |
| 35-44 | 36.0% | 39.0% | 24.0% |
| 45-54 | 30.1% | 40.6% | 28.8% |
| 55-64 | 26.5% | 42.3% | 30.6% |
| 65-74 | 25.1% | 44.0% | 30.0% |

^†^ Average proportion of normal weight, overweight and obesity for each age group among individuals aged 18-74, between 1998 and 2016

We then combined these weights to calculate the distribution of each age group by BMI group (see table S6.3).

Moreover, we estimated the BMI transitions from CALIBER for each age group and then we multiplied each of these transitions (table S6.4) with the corresponding proportions from table S6.3. In this way, we calculated the standardised transitions by BMI group (see figure 2 in the paper), after taking into consideration the age and BMI structure of the English population.

*Table S3.4.3: Age structure^†^ of the English population from HSE between 1998 and 2016, by BMI group*

| **Age group**  **(in years)** | **Proportion of normal weight** | **Proportion of overweight** | **Proportion of obesity** |
| --- | --- | --- | --- |
| 18-24 | 20.8% | 7.6% | 5.7% |
| 25-34 | 24.5% | 17.7% | 15.0% |
| 35-44 | 19.8% | 21.4% | 20.5% |
| 45-54 | 15.3% | 20.7% | 22.9% |
| 55-64 | 11.4% | 18.1% | 20.4% |
| 65-74 | 8.3% | 14.6% | 15.4% |
| Total | 100% | 100% | 100% |

^†^ Average proportion of each age group among individuals aged 18-74, between 1998 and 2016

*Table S3.4.4: Estimated transitions from CALIBER from normal weight, overweight and obese to other BMI groups at one, five and ten years, by age group*

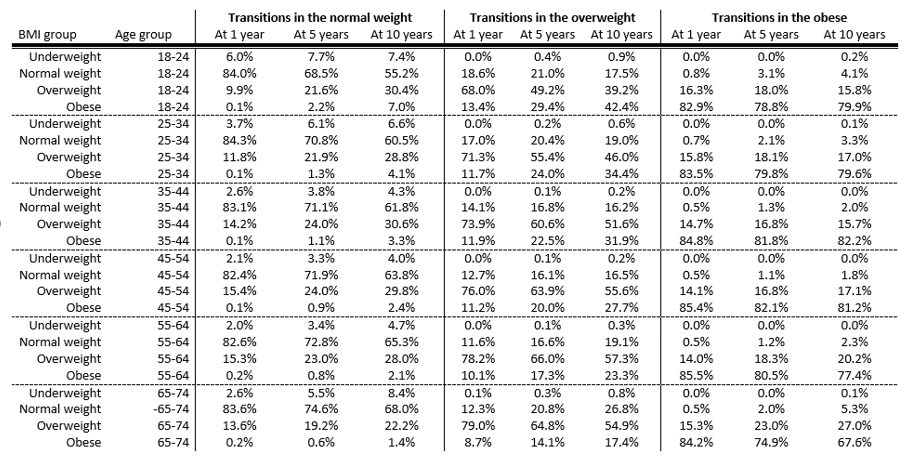

**3.5. Converting an odds ratio to a relative risk (for the online calculator)**

In figure 5, we calculated the absolute risk of transitioning to a higher BMI category over 10 years, by initial BMI category, age, sex and social deprivation non-parametrically. We wanted to do the same, this time accounting for ethnicity as well in our online calculator. If we did that non-parametrically, we would have near positivity issues; this means that in some strata defined by all potential categories of the sociodemographic factors, the number of individuals would be very small. For this reason, we used a semi-parametric approach: We calculated the (non-parametric) 10-year BMI transition by age, sex, index of multiple deprivation and current BMI overall is calculated in white individuals and then multiplied by the relative risk of transitions of ethnicity from figure 4. More specifically, we converted the corresponding odds ratios from 4 to relative risks using the formula from[1], see below

$$Relative risk= \frac{odds ratio}{1-p_{0}+(p_{0}\times odds ratio)}$$

where $p_{0}$ is the baseline risk, i.e. the risk in each cell of figure SXXX which is the absolute risk of transitioning to a higher BMI category over 10 years, by initial BMI category, age, sex and social deprivation non-parametrically in white individuals only.

References

[1] Grant RL. Converting an odds ratio to a range of plausible relative risks for better communication of research findings. BMJ 2014; 348:f7450.

**Section 4: Sensitivity analysis**

Figure S4.1: Absolute risks and odds ratios* of transitioning at ten years from normal weight to overweight or obese, from overweight to obese and from non-morbid to morbid obese, by age, sex, ethnicity, social deprivation and region (without adjusting for chronic diseases)

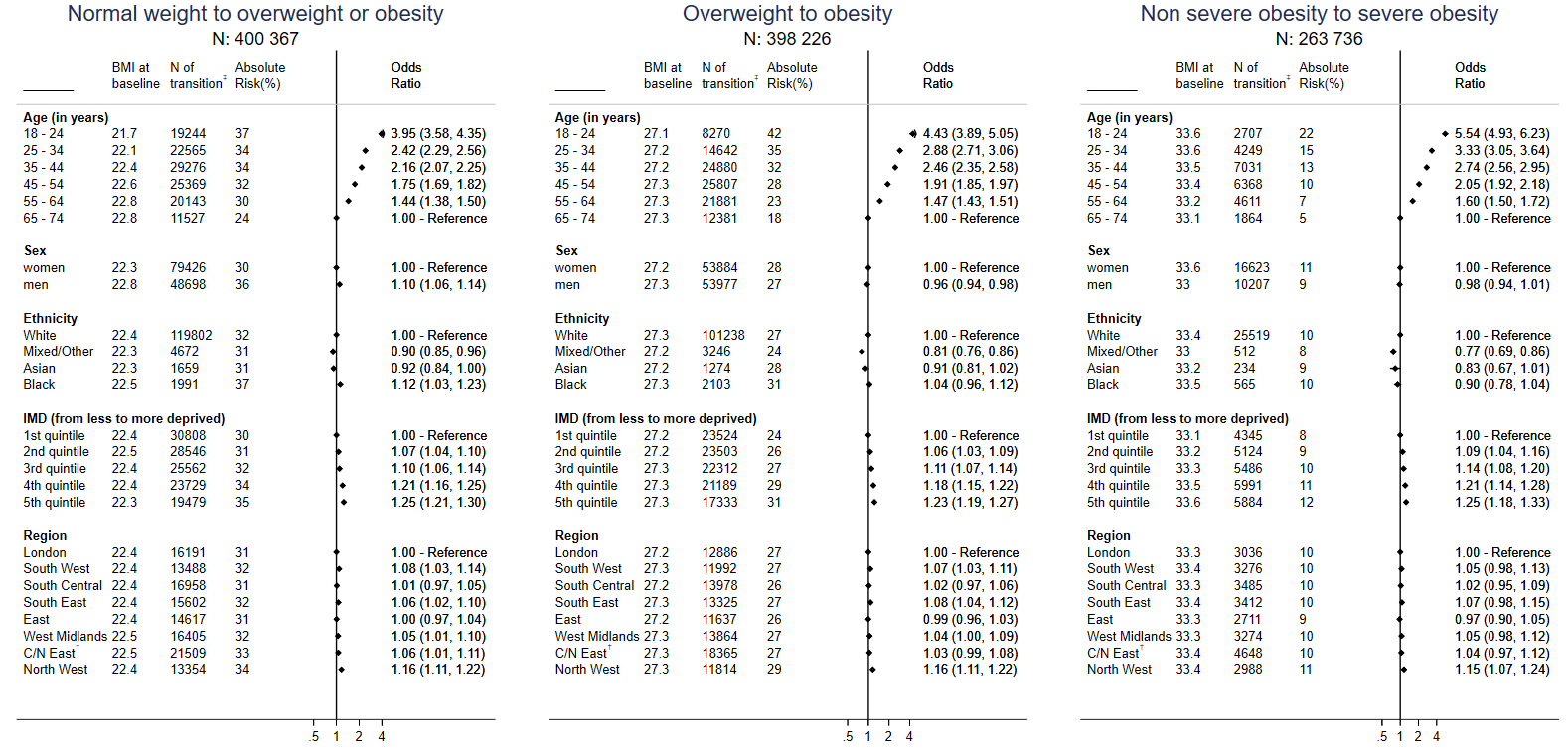

^*^Mutually adjusted for BMI (at baseline), age group, gender, Index of multiple deprivation (IMD – quintiles; in categories) and region

†Central/North East

‡N of individuals who transitioned to higher BMI categories

Figure S4.2: Absolute risks and odds ratios* of transitioning at ten years from normal weight to overweight or obese, from overweight to obese and from non-morbid to morbid obese, by age, sex, ethnicity, social deprivation and region, after excluding individuals with chronic diseases at baseline

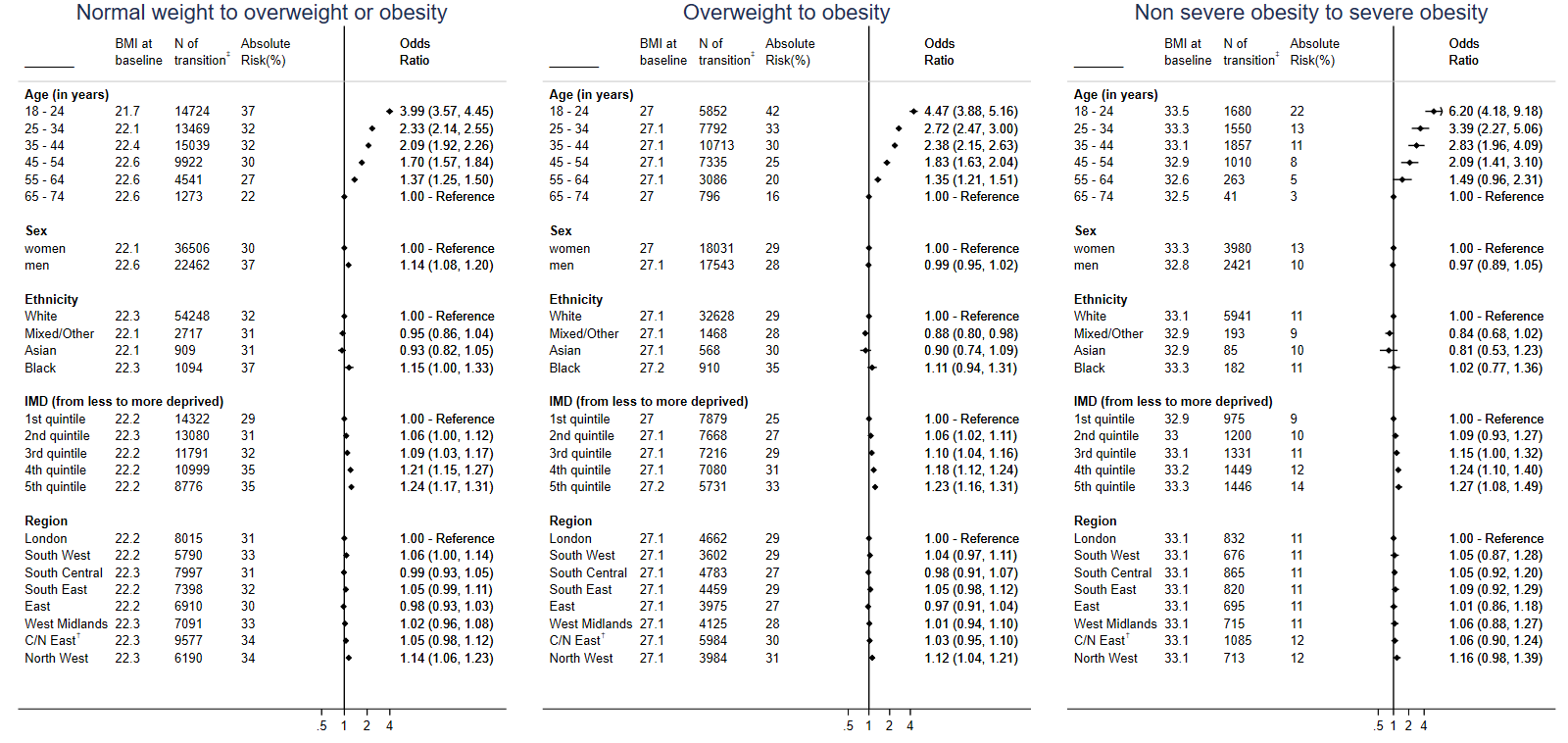

^*^Mutually adjusted for BMI (at baseline), age group, gender, Index of multiple deprivation (IMD – quintiles; in categories)

†Central/North East

‡N of individuals who transitioned to higher BMI categories

Figure S4.3: Absolute risks and odds ratios* of transitioning at ten years from normal weight to overweight or obese, from overweight to obese and from non-morbid to morbid obese, by age, sex, ethnicity, social deprivation and region, after excluding individuals with chronic diseases at baseline and follow-up

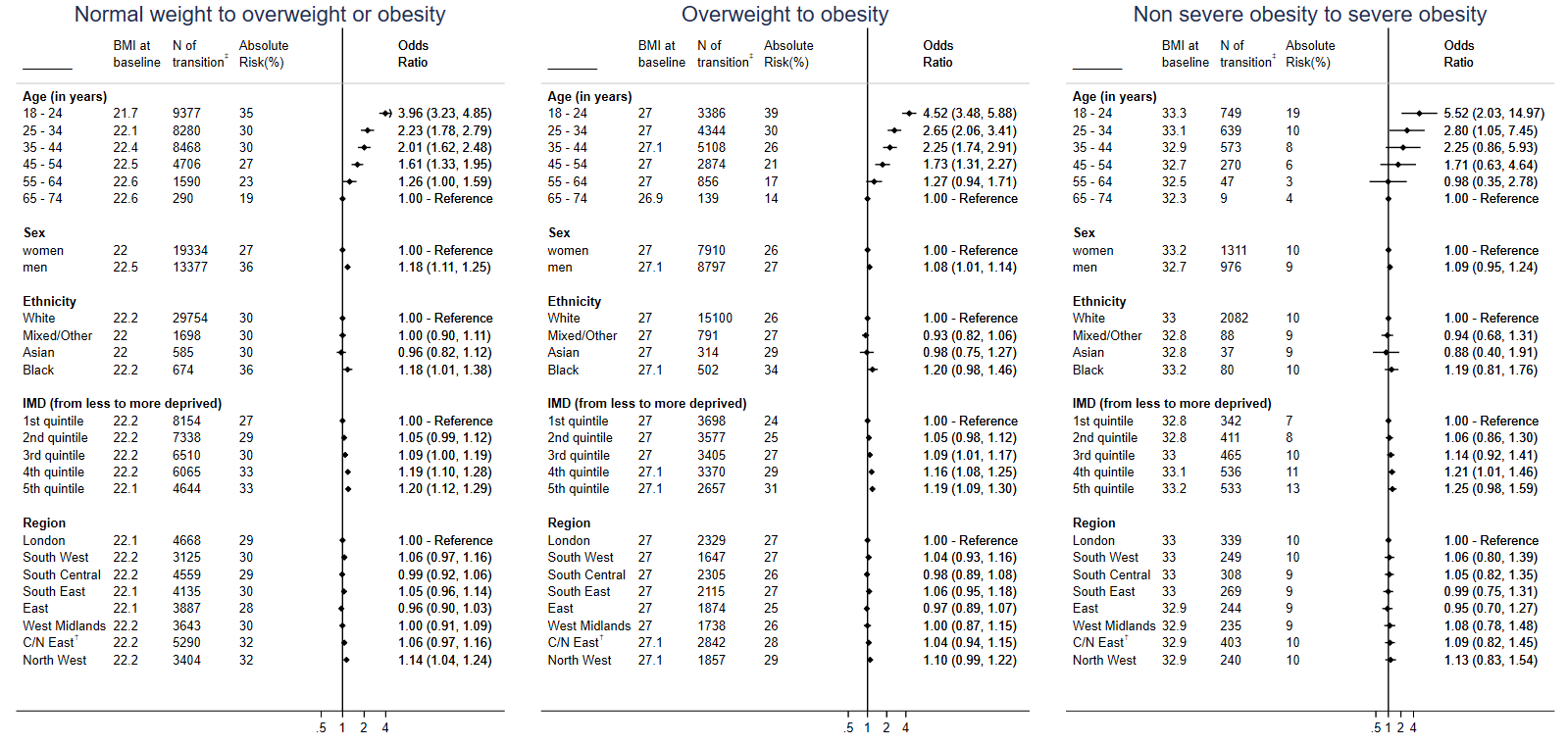

^*^Mutually adjusted for BMI (at baseline), age group, gender, Index of multiple deprivation (IMD – quintiles; in categories) and region

†Central/North East

‡N of individuals who transitioned to higher BMI categories

Figure S4.4: Absolute risks and odds ratios* of transitioning at ten years from normal weight to overweight or obese, from overweight to obese and from non-morbid to morbid obese, by age, sex, ethnicity, social deprivation and region. BMI change was calculated without delta adjustment

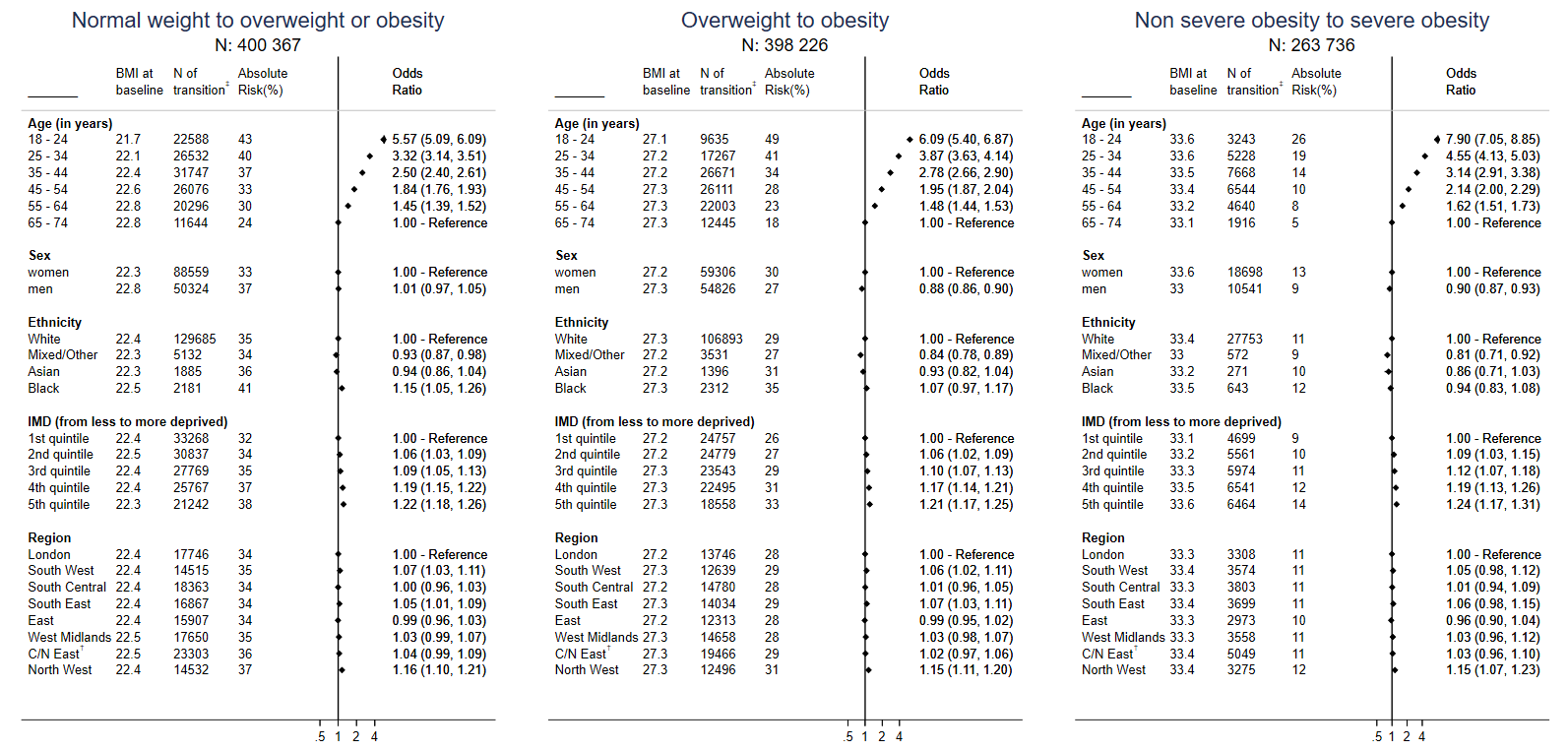

^*^Mutually adjusted for BMI (at baseline), age group, gender, Index of multiple deprivation (IMD – quintiles; in categories), region, prevalence of mental disease (bipolar disorder, anxiety, acute stress, affective disorder, phobia, schizophrenia and depression), prevalence of chronic diseases (cancer, dementia, rheumatoid arthritis, diabetes, COPD, renal disease, gout, HIV, chronic kidney disease, inflammatory bowel disease, systemic lupus erythematosus and cardiovascular diseases), prevalence of hypertension, use of diuretics

†Central/North East

‡N of individuals who transitioned to higher BMI categories

Figure S4.5: Absolute risks and odds ratios* of remaining obese at ten years, by age, sex, ethnicity, social deprivation and region. Results from the complete case analysis

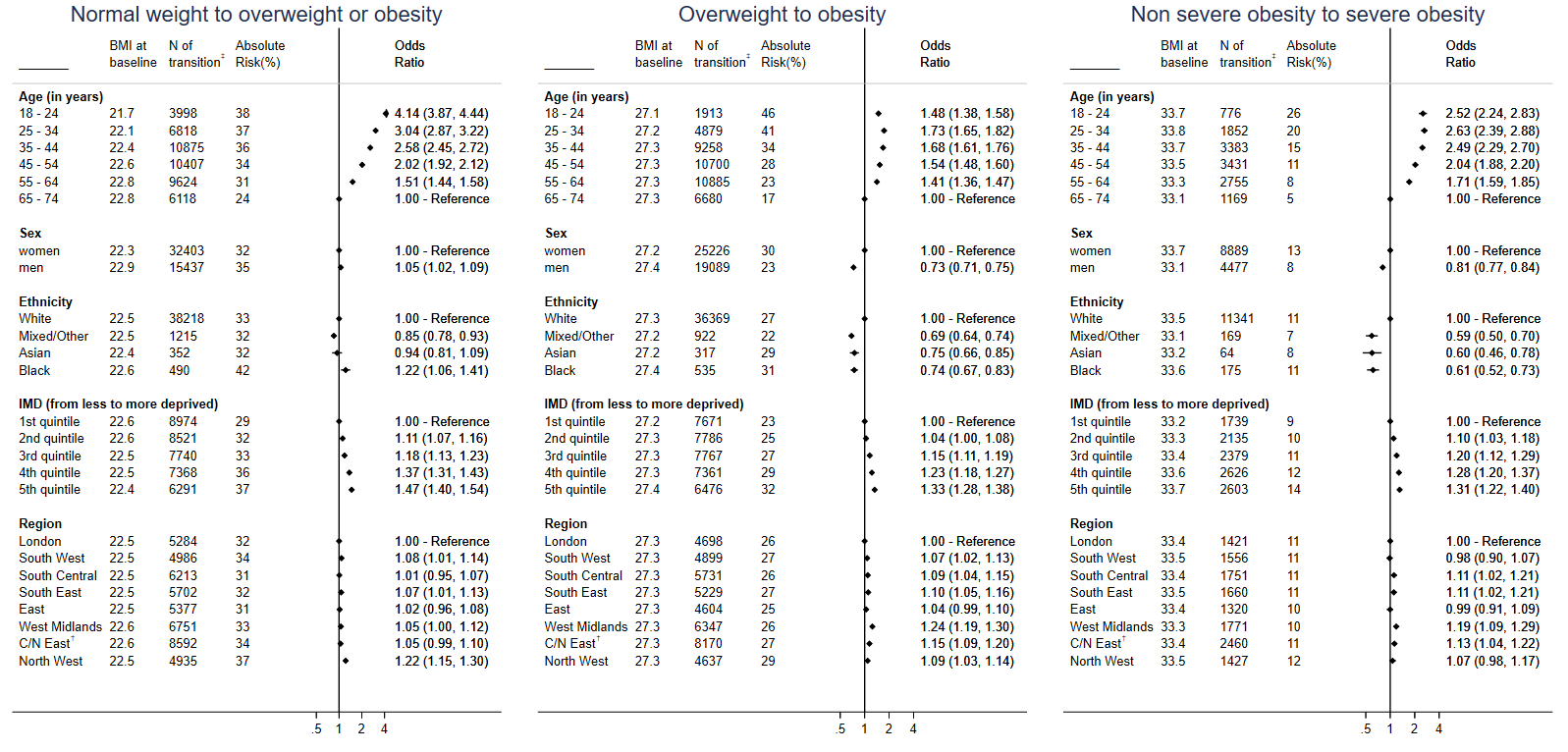

^*^Mutually adjusted for BMI (at baseline), age group, gender, Index of multiple deprivation (IMD – quintiles; in categories), region, prevalence of mental disease (bipolar disorder, anxiety, acute stress, affective disorder, phobia, schizophrenia and depression), prevalence of chronic diseases (cancer, dementia, rheumatoid arthritis, diabetes, COPD, renal disease, gout, HIV, chronic kidney disease, inflammatory bowel disease, systemic lupus erythematosus and cardiovascular diseases), prevalence of hypertension, use of diuretics

†Central/North East

‡N of individuals who transitioned to higher BMI categorie
